# Antimicrobial-resistant *E. coli* in human, animal and environmental reservoirs in rural Bangladeshi households with young children

**DOI:** 10.64898/2026.06.16.26355831

**Authors:** Sumaiya Tazin, Md. Sakib Hossain, Ashrin Haque, Md. Hajbiur Rahman, Tahani Tabassum, Amanta Rahman, Claire Anderson, Suhi Hanif, Gabriella Barratt Heitmann, Md. Rana Miah, Afsana Yeamin, Farjana Jahan, Abul Kasham Shoab, Mahbubur Rahman, Zahid Hayat Mahmud, Jade Benjamin-Chung, Ayse Ercumen

**Author notes:** Corresponding Author: Jordan Hall Addition, Raleigh, North Carolina, 27695, United States.

## Abstract

In low-income countries, ESBL-producing *Escherichia coli* (ESBL-EC) is frequently detected in humans, animals and household environments, indicating widespread exposure to antimicrobial resistance (AMR). Established risk factors such as antibiotic use do not explain the high community carriage of AMR in all settings; identifying the dominant exposure pathways can inform interventions against AMR. We aimed to investigate (i) animal-human-environment sharing of AMR by assessing associations between the abundance of ESBL-EC in the household environment, domestic animal feces and young children’s stool and (ii) household factors associated with ESBL-EC abundance in these reservoirs. We enrolled 112 households from the CRADLE trial in rural Bangladesh. We enumerated ESBL-EC in drinking water, food, child hand rinses, outdoor soil, indoor floor swabs, chicken and cow feces, and stool from children aged 6 months. We recorded indicators of sanitation, animal ownership/management, human and animal antibiotic use, and child exposure behaviors using structured questionnaires and spot checks. The highest prevalence of ESBL-EC was in child stool (95.6%) and animal feces (82.3-96.9%), followed by soil (48.2%) and floors (36.6%); < 10% of food, child hands and drinking water harbored ESBL-EC. The abundance of ESBL-EC in child stool was not associated with its abundance in any sampled matrix; the abundance in chicken but not cow feces showed positive correlations with soil, floors, child hands, and drinking water (correlation coefficients: 0.19-0.39, p-values < 0.05). Higher-quality latrines (improved, pour-flush, with slab) were associated with lower ESBL-EC abundance across matrices; unsafe animal management (animals roaming or spending the night inside the home) was associated with higher abundance. Child antibiotic use and exposure behaviors (soil ingestion, time spent on floor) were not associated with ESBL-EC abundance in child stool. We observed high AMR colonization among young children and domestic animals in rural Bangladesh not explained by traditional fecal-oral exposure pathways. Future studies should explore additional pathways and assess whether sanitation and animal management improvements can reduce AMR.

## Introduction

Antimicrobial resistance (AMR) is a growing public health concern, with a particularly high burden in low- and middle-income countries. Extended-spectrum β-lactamase (ESBL) producing organisms are of particular concern; these antimicrobial-resistant bacteria can break down beta-lactam antibiotics, including penicillins, most cephalosporins (1^st^, 2^nd,^ and 3^rd^ generations) and monobactams, making infections difficult to treat. With limited treatment options available, resistant infections often require last-resort antibiotics like carbapenems, increasing the risk of further resistance development. Among six world regions, individuals from Southeast Asia had the highest prevalence (27%) of gut colonization with ESBL-producing *E. coli.*^1^

Drivers of AMR in low-income countries include overuse of antibiotics, both for human and animal use.^2^ However, global literature suggests that environmental exposures are a stronger contributor to the spread of AMR than antibiotic consumption.^3–5^ In settings where fecal waste is not safely isolated from the environment, antimicrobial-resistant bacteria can be transmitted between human or animal hosts and environmental reservoirs, and environmental matrices (e.g. waterbodies, soils) can amplify resistance by enabling genetic exchange between fecal and native microorganisms.^6–8^ In low-income countries, ESBL-producing *E. coli* has been detected in surface waters,^9,10^ drinking water supplies,^11^ food,^12–14^ domestic soils,^15^ and indoor household floors,^16–18^ highlighting its potential for widespread environmental transmission.

The complexity of AMR transmission is increased by domestic animals that typically share living space with household members in low- and middle-income countries.^19^ Therapeutic and sub-therapeutic use of antimicrobials is common in backyard or small-scale animal husbandry in low-income countries.^20,21^ ESBL-producing *E. coli* is frequently detected in feces of chickens^20,22,23^ and cattle^24^ raised in these settings. Unsafely managed animal fecal waste can contribute to contamination of the domestic environment with fecal organisms,^25^ including antimicrobial-resistant bacteria.^17^ Consequently, ESBL-producing *E. coli* may transmit between animal and human hosts via multiple environmental pathways.^26^ Antibiotic consumption in animals has been linked with AMR in human pathogens,^27^ and studies have shown overlapping resistomes between humans and domestic animals in low-income countries.^28^ Soil in the domestic environment (e.g. yards, indoor soil floors) may be an environmental reservoir of particular concern because it harbors a high density of native microorganisms, is a repository for animal fecal waste, and may therefore facilitate AMR transmission between domestic animals and humans sharing living spaces.^29^

Children are particularly vulnerable to environmental exposure to fecal organisms because of age-specific behaviors such as soil ingestion (geophagia)^30^ and mouthing of contaminated hands and objects.^31,32^ A study in rural Bangladesh reported that 74% of healthy infants <12 months were colonized with ESBL-producing *E. coli*.^33^ However, established risk factors such as history of antibiotic use and hospitalization, or other factors such as the infant’s gender, religion or feeding practices were not associated with colonization.^33^ A recent study in Bangladesh found that, among newborns, 72% were colonized with ESBL-producing bacteria, and 54% were colonized with carbapenem-resistant bacteria within 2-7 days of birth, highlighting an alarmingly early exposure to AMR organisms.^34^ Among healthy infants in Bangladesh, whole genome sequencing indicated that transmission from colonized mothers and drinking water exposures may contribute to child colonization with ESBL-producing *E. coli*, ^35^ indicating that environmental exposures within the home can play a role. Potential AMR transmission along other fecal-oral pathways in the domestic environment (e.g., food stored in the home, hands, soil, objects, surfaces) has not been investigated. Importantly, some of these matrices (e.g. soil) have substantially higher levels of fecal contamination than drinking water,^25^ and non-waterborne pathways (e.g. food, child hands, soil, objects) are dominant sources of *E. coli* ingestion among young children;^31^ these pathways could be overlooked drivers of AMR transmission.

Although WHO recommends environmental surveillance for ESBL-producing *E. coli* to monitor the environmental spread of AMR,^36^ current surveillance is primarily focused on waterbodies.^37^ Few studies have conducted integrated AMR surveillance across human, animal and environmental reservoirs in low-income countries to identify pathways of AMR exposure, particularly among children.^38,39^ Similarly, studies have assessed some risk factors for exposure (e.g. antibiotic use, animal ownership) but how multiple, concurrent drivers - including environmental, animal and household-level exposures - interact to shape child colonization with AMR has not been examined. Understanding the sources, transmission pathways, and risk factors associated with child colonization with ESBL-producing *E. coli* in the community setting is crucial for developing effective infection control strategies and mitigating public health impacts.

In this study, we aimed to explore animal-human-environment sharing of AMR by assessing associations between the prevalence and abundance of ESBL-producing *E. coli* in the household environment (drinking water, food, outdoor soil, indoor floors), domestic animal feces (chickens, cows) and stool samples from young children. Further, we assessed household and environmental factors associated with the prevalence and abundance of ESBL-producing *E. coli* in the home environment and among domestic animals and children.

## Methods

### Study design and setting

We used data from the Cement-based flooRs AnD chiLd hEalth (CRADLE) trial (NCT05372068). CRADLE is a randomized controlled trial in rural villages of Sirajganj and Tangail districts in Bangladesh that aims to assess the health effects of replacing soil floors with concrete floors; the trial design and specifics have been previously published.^40^ The study villages are adjacent to Jamuna, a flood prone river, and experience flooding frequently.^41^ Animal husbandry, particularly cattle rearing, is common in the study area, and humans and animal live in close proximity.^42^

In brief, the CRADLE trial enrolled pregnant women, installed concrete floors in intervention households when the birth cohort (i.e., index children) was in utero and measured environmental and child health outcomes at index child ages 3, 6, 12, 18, and 24 months. The current study used data from the trial’s baseline and 6-month follow-up data collection. ESBL-producing *E. coli* is one of the secondary outcomes of the trial and was measured among a random subset of trial participants at the 6-month follow-up. Socio-demographic indicators (e.g. mother’s age and education, household size, assets) were recorded at baseline.^40^

### Data collection

Trained field staff from the International Centre for Diarrhoeal Disease Research, Bangladesh (icddr,b) aimed to visit a random subset of 110 households (55 intervention, 55 control) enrolled in the CRADLE trial for an environmental sub-study between May 2024 and August 2025. In each household, they conducted a structured questionnaire and spot check observations to record demographics, sanitation practices, animal ownership and management practices, antibiotic use among children and domestic animals, and index child activities with respect to animal and environmental exposures.

### Sample collection

In each household, field staff trained in sterile technique collected samples of index child stool and domestic animal feces, as well as samples from the domestic environment, including stored drinking water, prepared food (preferably rice), child hand rinse, courtyard soil, and floor swab from the child’s sleeping area. All samples were transported to the Laboratory of Environmental Health at icddr,b at 4-10°C using coolers with ice and preserved overnight at 4°C.

#### Index child stool

Field staff collected stool samples from the index child aged 6 months old. A stool collection kit containing aluminum foil was provided to the caregiver the day before sampling. The caregiver was asked to collect the child’s stool the next morning on the foil or on a diaper if unable to collect the stool on the foil. The caregiver was directed to scoop the stool from the top of the pile as soon as possible after defecation into the sterile stool collection tube using the sterile spoon attached to the lid of the tube. They were directed to keep the tube away from sunlight and to remember the exact time of defecation. At least 15 mL of stool was collected for the index child. Field staff picked up the collection tubes the next day.

#### Chicken and cow feces

Field staff collected chicken and cow feces in each household if fresh feces were available. Feces defecated since dawn/early morning on the day of sampling was defined as fresh. Staff prioritized collecting feces from the same room as the floor samples if available. Otherwise, they collected feces samples from either inside the house where the index child lived or from the courtyard/compound area (compound refers to multiple households from an extended family with a shared courtyard). If feces were not found within the living space, samples were collected from where households collected and stored animal feces (e.g. bucket) if fresh feces had been added to the pile since dawn/early morning. The feces were scooped with a sterile spoon to fill a 40-mL sterile plastic tube (one tube for chicken feces, one tube for cow feces). The tubes were placed in individual Ziploc bags to prevent cross-contamination with other samples.

#### Stored drinking water

Field staff asked participants for a glass of drinking water from their storage container or the source the same way they would give it to the index child or other children <5 years. Water was poured into a sterile Whirlpak bag (Nasco, Modesto, CA) to collect approximately 150 mL of sample.

#### Prepared food

Respondents were asked for food items stored for the consumption of the index child (e.g. rice, khichuri – lentil and rice stew, or suji – semolina porridge). If the index child was not yet eating solid food or if stored food for the index child was not available, respondents were asked for food samples stored for other children <5 years, and otherwise for adults. The food was scooped with a sterile spoon to fill a 40-mL sterile plastic tube.

#### Child hand rinse

Hand rinses were collected from index children or the youngest child <5 years if the index child was unavailable. The caregiver was asked to place one hand of the child at a time into a sterile Whirlpak bag pre-filled with 250 mL of sterile DI water. The hand was massaged from outside the bag for 15 seconds and shaken horizontally for 15 seconds. The same procedure was repeated for the other hand within the same bag, and the rinse water was saved.

#### Courtyard soil

Field staff identified a courtyard area immediately adjacent to the entrance of the household where the index child lived. They placed an ethanol- and bleach-sterilized stencil on a 30 cm x 30 cm area and collected soil samples by scraping the area inside the stencil with a sterile plastic scoop to fill a sterile 40-mL tube. The tube was placed in a Ziploc bag to prevent cross-contamination with other samples.

#### Floor swab

Field staff identified the nearest area next to the head of the bed where the index child usually slept and placed an ethanol- and bleach-sterilized stencil on a 50 cm x 50 cm area. They swabbed the area inside the stencil once from side to side and once from top to bottom in S-shaped motion with a Hydrated PolySponge (Nasco, Modesto, CA) and placed the sponge in a sterile Whirlpak bag.

#### Quality control for environmental samples

10% field blanks and duplicates for environmental samples were collected for quality control. For water samples, blanks were collected by transferring 100 mL of sterile DI water into the sterile Whirlpak in the same way the water samples were collected. For hand rinse samples, blanks were collected by performing the massaging and shaking steps without immersing a child hand in the prefilled Whirlpak. For swab samples, blanks were collected by removing the sponge from the Whirlpak bag, then putting it back inside the Whirlpak without swabbing a surface. For swab samples, duplicates were collected by swabbing the nearest area to the initial sample where it was possible to fully swab a 50 cm x 50 cm area.

### Sample analysis

Samples were processed within 24 hours of collection. Environmental samples (water, hand rinse, food, courtyard soil, floor swabs) were processed with membrane filtration. Solid samples were homogenized and floor swabs were eluted prior to membrane filtration. For solid samples (food, soil), a 10 g aliquot was homogenized with 90 mL of sterile saline solution. A separate 5 g aliquot of food and soil samples was oven-dried at 110°C for 24 hours to obtain the dry weight. For floor swab samples, lab staff added 100 mL of sterile DI water to the Whirlpak containing the sponge, massaged the sponge from outside the bag for 15 sec, and then swirled the bag for an additional 15 sec. The liquid was decanted into a new sterile Whirlpak, then the procedure was repeated two more times to yield a total of 300 mL eluate. The following volumes were used for membrane filtration based on feasibility of filtering without clogging the filter: 100 mL of drinking water, 100 mL of child hand rinse, 10 mL of soil homogenate, 10 mL of food homogenate, and 20 mL of floor swab eluate.

Animal and child fecal samples were analyzed using the drop plate technique. 10 g of sample was homogenized with 90 mL of sterile saline solution, and serial 1:10 dilutions were performed as needed to obtain colonies in the countable range. If 10 g of the sample was not available, the maximum available amount was used while maintaining the same ratio of sample to saline (e.g., 5 g of sample homogenized with 45 mL of saline for a 1:10 dilution). The following volumes of the homogenate were plated: 50 µL for chicken and cow feces, 25 µL for child stool.

ESBL-producing *E. coli* was enumerated using CHROMagar™ ESBL agar. Plates were incubated at 37 °C for 18-24 hrs, and presumptive ESBL-producing *E. coli* colonies were enumerated based on colony appearance. 10% lab blanks were processed using sterile DI water.

### Ethical considerations

The study was approved by the Institutional Review Board of Stanford University (63990) and the Ethical Review Committee of the icddr,b (PR-22069). Female caregivers of the child <2 years provided written informed consent in Bengali.

### Statistical analysis

#### Outcomes

For each sample type, we calculated the prevalence and abundance of ESBL-producing *E. coli.* We defined abundance as the log10-transformed colony forming units (CFU) per reporting unit. The reporting units are per 100 mL for water samples, per two hands for child hand rinses, per dry gram for food and soil samples, per unit floor area for swab samples, and per gram for animal feces and child stool samples. Non-detects were imputed with half the lower detection limit prior to taking the logarithm. Lower detection limits varied by sample type, dilution factor used and moisture content (Table S1).

#### Associations between ESBL-producing *E. coli* in domestic environment, animal feces and child stool

We generated a correlation matrix using pairwise comparisons of ESBL-producing *E. coli* abundance across all sample types. Spearman’s rank correlation coefficients (ρ) were calculated to account for non-normal distributions. A p-value <0.05 was considered a statistically significant correlation. Additionally, we used generalized linear models to assess adjusted associations between the log10-transformed CFU of ESBL-producing *E. coli* in environmental samples and animal feces vs. the log10-transformed CFU of ESBL-producing *E. coli* in child stool; for sample types with low levels of contamination (>90% non-detects), we used the binary prevalence of ESBL-producing *E. coli* as the independent variable. The models used Gaussian error distribution and robust errors to account for geographical clustering of households enrolled in the CRADLE trial. The models adjusted for potential socio-demographic confounders (child age and sex, mother’s age and education, number of children <18 years in the household, number of individuals living in the compound, and asset-based wealth index), study arm (intervention vs. control) and whether children aged 3-8 years open defecated, poultry were kept inside the home at night, and domestic animals were given antibiotics. We expect these factors to be independent predictors of both the outcome variable (ESBL-producing *E. coli* in children) and exposure variables in this analysis (ESBL-producing *E. coli* in the environment and animals).

#### Drivers of ESBL-producing *E. coli* in domestic environment, animal feces and child stool

This analysis used the log10-transformed CFU of ESBL-producing *E. coli* in each sample type as outcome variables and a set of household factors as exposure variables. We excluded sample types with low levels of contamination (>90% non-detects) from this analysis. We grouped outcomes within the following domains: (O1) environment, (O2) animals and (O3) children, and exposures within the following domains: (E1) sanitation, (E2) animal ownership and management, (E3) antibiotic use (including use by index child, index mother, domestic animals), and (E4) child exposure behaviors (birth location, soil ingestion, contact with floors, animal contact) (Fig 1). Within each domain, we considered several self-reported and observed variables; we excluded variables that did not have sufficient variation (<5% of responses in one category) without our study sample (Table S2). For each outcome domain, we assessed relationships with the exposure variables that are upstream of the outcome (Fig1, Table S3). Models used generalized linear models with robust standard errors. Models for continuous outcomes used a Gaussian error distribution, and models for binary outcomes used a Poisson error distribution with a log link. Given the complexity of the causal relationships within and between the exposure domains and our small sample size, models explored one exposure variable at a time, controlling for mother’s age and education, number of children <18 years in the household, number of individuals living in the compound, wealth index and study arm. Models for child colonization also controlled for child age and sex.

**Fig 1.**
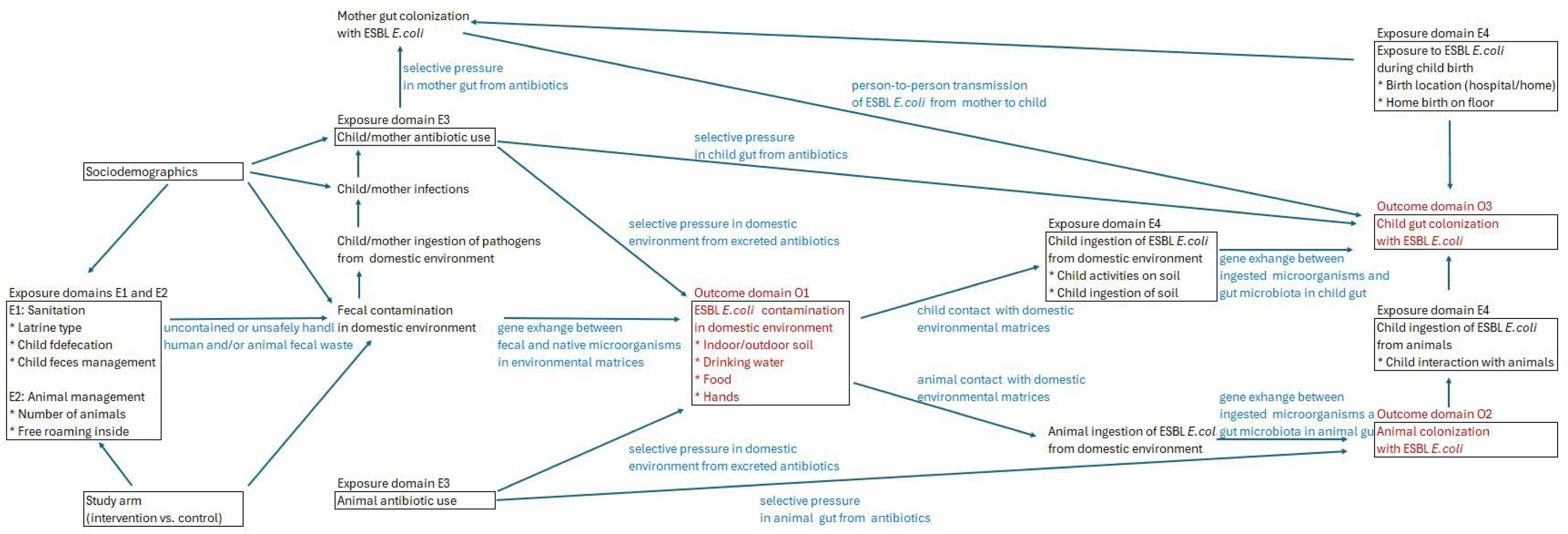
Hypothesized relationships between household factors and ESBL-producing *E. coli* in the domestic environment, animal feces, and child stool

## Results

### Enrollment

A total of 112 households (113 index children including a pair of twins) were enrolled in the environmental sub-study of the CRADLE trial between May 2024 and August 2025. Staff collected 113 samples of index child stool and index child hand rinses, 112 samples of floor swabs, courtyard soil and drinking water, 103 food samples, 65 chicken feces samples, and 62 cow feces samples (Table 1). The lower number of food and animal feces samples were due to lower availability (e.g., no stored food present, no animals in compound, no fresh animal feces available).

**Table 1.** ESBL-producing *E. coli* prevalence and abundance by sample type in drinking water, food, child hands, floor swabs, courtyard soil, chicken feces, cow feces, and child stool.

| Sample type | N | n Positive | Prevalence % | Mean log<sub>10</sub>-CFU | SD log<sub>10</sub> CFU | Reporting unit |
| --- | --- | --- | --- | --- | --- | --- |
| Drinking water | 112 | 1 | 0.9 | -0.28 | 0.17 | Per 100 mL |
| Food | 103 | 6 | 5.8 | -0.33 | 0.57 | Per 1 dry g |
| Child hands | 113 | 10 | 8.8 | 0.17 | 0.28 | Per 2 hands |
| Floor swabs | 112 | 41 | 36.6 | 1.68 | 1.20 | Per 0.25 m<sup>2</sup> |
| Courtyard soil | 112 | 54 | 48.2 | 1.04 | 1.54 | Per 1 dry g |
| Chicken feces | 65 | 63 | 96.9 | 5.42 | 1.32 | Per 1 g |
| Cow feces | 62 | 51 | 82.3 | 3.61 | 1.39 | Per 1 g |
| Child stool | 113 | 108 | 95.6 | 6.60 | 1.65 | Per 1 g |

CFU: colony-forming unit, SD: Standard deviation

### Household characteristics

#### Socio-demographics

Among 112 households enrolled in environmental sub-study, the mean household size was 5, the mean age of the mother of the index child was 24 years, and the mothers had on average 7 years of schooling (Table S4). Approximately 98% houses had finished walls and electricity, 31% had a refrigerator, and 12% had a television. All households used tubewells as their main drinking water source.

#### Sanitation

Latrine access was high, where 85.7% (96) of households had an improved latrine as defined by the Joint Monitoring Program (Table S4). Open defecation was not reported among older children and adults while almost all children <3 years (98.2%, n = 110) and 20.3% (15) of children 3-8 years practiced open defecation daily. Safe defecation (in a potty, nappy, latrine) was reported for 43.8% (49) of index children, and 23.4% (26) of households reported disposing of child feces in a latrine.

#### Animal ownership and management

More than three quarters (77.7%, n = 87) of households owned domestic animals (68.8% chickens, 32.1% goats/sheep, 54.5% cows/buffalo), and 85.7% (96) of households were in a compound that kept animals (79.5% chickens, 42.0% goats/sheep, 54.5% cows/buffalo) (Table S4). The mean number of animals was 9.9 per household and 20.1 per compound. Chickens were reported to roam freely (sometimes/always) inside the house in 91.7% (103) and goats/sheep in 32.1% (36) of households, while cows/buffalo almost never roamed inside the house. Chickens were reported to roam freely (sometimes/always) in the compound in 73.2% (82), goats/sheep in 97.3% (109) and cows/buffalo in 38.4% (43) of households. In the room where the index child slept, staff observed animal feces in 48.2% (54) of households and animals in 36.6% (41) of households. In the food storage area, staff observed flies in 36% (37) of households but rarely observed animals or animal feces.

#### Human and animal antibiotic use

More than two thirds (67.9%, n = 76) of index children and 7.1% (8) of their mothers used antibiotics within the last six months (Table S4). The most reported antibiotics were beta-lactams (cefixime) and macrolides (azithromycin). A quarter (25.9%, n = 29) of index children and 1.8% (2) mothers reported using beta-lactam antibiotics in the last six months. Among 96 households in the compounds that kept animals, 22.9% (22) reported using antibiotics for their animals in the last six months. Most respondents did not know the name or class of the antibiotic used for animals.

#### Child exposure behaviors

Of 113 index children, 7.1% (8) were delivered in the hospital, while the remaining 92.9% (104) were delivered at home, and 48.2% (54) were delivered on the floor of the home (Table S4). Index children slept on the floor every day in 15.2% (17) of households and played on the floor every day in 62.5% (70) of households. Caregivers reported that 18.9% (21) of index children had eaten soil inside the home and 35.1% (39) had eaten soil outside the home within the last two days.

### ESBL-producing E. coli prevalence and abundance

Almost all index children (96%, n=108) had ESBL-producing *E. coli* in stool with a mean abundance of 6.6 log10-CFU/g (Table 1). Among all sample types, we observed the highest prevalence of ESBL-producing *E. coli* in chicken feces (97%, n = 63) with a mean abundance of 5.2 log10-CFU/g, while 82% (51) of cow feces harbored ESBL-producing *E. coli* with a mean abundance of 3.6 log10-CFU/g (Table 1). Among environmental samples, we observed the highest prevalence of ESBL-producing *E. coli* in courtyard soil (48%, n = 54, mean = 1 log10-CFU/dry g) and floor swabs (36%, n = 44, mean = 1.7 log10-CFU/0.25 m^2^) (Table 1). Less than 10% of child hand rinse and food samples and 1% of drinking water samples harbored ESBL-producing *E. coli* (Table 1).

### Relationships between abundance of ESBL-producing E. coli across sample types

The abundance of ESBL-producing *E. coli* in child stool was not significantly correlated with the abundance of ESBL-producing *E. coli* in animal feces samples or any type of environmental sample (Tables 2 and 3). The abundance in chicken feces, but not cow feces, was positively correlated with the abundance in courtyard soil (ρ = 0.27, p-value = 0.03) and in floor swabs (ρ = 0.35, p-value <0.005), while the abundance in courtyard soil and abundance in floor swabs were also positively correlated (ρ = 0.39, p-value <0.005) (Table 2). In turn, the abundance in courtyard soil was positively correlated with the abundance on child hands (ρ = 0.19, p-value = 0.04), while the abundance in floors swabs was correlated with the abundance on child hands (ρ = 0.19, p-value = 0.05) and in drinking water (ρ = 0.19, p-value = 0.05) (Table 2). The abundance on child hands and in drinking water were also positively correlated.

**Table 2.** Pairwise Spearman correlation coefficients for log10-CFU of ESBL-producing *E. coli* across sample types. Values in parentheses denote p-values. Red cells denote positive correlation, blue cells denote negative correlation, bolded font denotes significant correlation with p-value<0.05.

| Samples | Drinking water | Stored food | Child hands | Floor swabs | Courtyard soil | Chicken feces | Cow feces | Child stool |
| --- | --- | --- | --- | --- | --- | --- | --- | --- |
| Drinking water | 1 | 0.16 (0.11) | **0.31 (<0.005)** | **0.19 (0.05)** | 0.16 (0.10) | 0.12 (0.33) | --<sup>a</sup> | -0.08 (0.42) |
| Food | 0.16 (0.11) | 1 | 0.01 (0.93) | -0.07 (0.46) | -0.18 (0.07) | -0.02 (0.87) | -0.04 (0.75) | 0 (0.97) |
| Child hands | **0.31 (<0.005)** | 0.01 (0.93) | 1 | **0.19 (0.05)** | **0.19 (0.04)** | 0.13 (0.32) | 0.03 (0.80) | 0.12 (0.21) |
| Floor swabs | **0.19 (0.05)** | -0.07 (0.46) | **0.19 (0.05)** | 1 | **0.39 (<0.005)** | **0.35 (<0.005)** | -0.06 (0.65) | 0.11 (0.26) |
| Courtyard soil | 0.16 (0.10) | -0.18 (0.07) | **0.19 (0.04)** | **0.39 (<0.005)** | 1 | **0.27 (0.03)** | 0.12 (0.34) | -0.08 (0.42) |
| Chicken feces | 0.12 (0.33) | -0.02 (0.87) | 0.13 (0.32) | **0.35 (<0.005)** | **0.27 (0.03)** | 1 | 0.24 (0.11) | -0.06 (0.65) |
| Cow feces | --<sup>a</sup> | -0.04 (0.75) | 0.03 (0.80) | -0.06 (0.65) | 0.12 (0.34) | 0.24 (0.11) | 1 | -0.02 (0.85) |
| Child stool | -0.08 (0.42) | 0 (0.97) | 0.12 (0.21) | 0.11 (0.26) | -0.08 (0.42) | -0.06 (0.65) | -0.02 (0.85) | 1 |

CFU: colony-forming unit
<sup>a</sup> Correlation between drinking water and cow feces was not estimable due to insufficient overlapping observations and lack of variance.

**Table 3.** Associations between log10-CFU of ESBL-producing *E. coli* in child stool vs. log10-CFU of ESBL-producing *E. coli* in floor swabs, courtyard soil, and animal feces and binary prevalence of ESBL-producing *E. coli* in drinking water, food and child hand rinses.

|  | N | Unadjusted |  | Adjusted<sup>a</sup> |  |
| --- | --- | --- | --- | --- | --- |
|  |  | ΔLog10 (95% CI) | p-value | ΔLog10 (95% CI) | p-value |
| Floor swab | 111 | 0.09 (-0.14, 0.32) | 0.42 | 0.13 (-0.13, 0.39) | 0.33 |
| Courtyard soil | 111 | -0.09 (-0.32, 0.14) | 0.43 | -0.17 (-0.37, 0.03) | 0.09 |
| Cow feces | 62 | -0.06 (-0.38, 0.25) | 0.69 | -0.09 (-0.37, 0.19) | 0.52 |
| Chicken feces | 65 | 0.05 (-0.23, 0.33) | 0.72 | -0.03 (-0.25, 0.18) | 0.77 |
| Drinking water | 112 | -0.72 (-0.99, -0.44) | **<0.005** | 0.20 (-0.99, 1.39) | 0.74 |
| Food | 103 | -0.02 (-0.62, 0.58) | 0.94 | 0.09 (-0.66, 0.84) | 0.82 |
| Child hands | 113 | 0.43 (-0.69, 1.56) | 0.45 | 0.16 (-0.80, 1.13) | 0.74 |

CFU: colony-forming unit, CI: confidence interval.
ΔLog10-CFU: The difference in log10-CFU between a comparison group and the reference group
<sup>a</sup> Adjusted analyses controlled for potential socio-demographic confounders (child age and sex, mother's age and education, number of children <18 years in the household, number of individuals living in the compound, and asset-based wealth index), study arm (intervention vs. control) and whether children aged 3-8 years open defecated, poultry were kept inside the home at night, and domestic animals were given antibiotics. For cow and chicken feces, we did not adjust for whether domestic animals were given antibiotics, and for chicken feces, we did not adjust for whether poultry were kept inside the home at night because we expect these variables to lie on the causal pathway. Estimates with p-values<0.05 are denoted in bold.

### Associations between abundance of ESBL-producing E. coli and household factors

#### Child stool

In adjusted analyses controlling for household socio-demographics, study arm, child age and child sex, children had approximately 1-log higher ESBL-producing *E. coli* in stool if index child feces were disposed of in the latrine as opposed to left there or disposed of in bush/garbage (ΔLog10 = 0.90 (0.36, 1.44)), if there were flies in the food storage area (ΔLog10 = 0.70 (0.09, 1.30)), if chickens stayed inside the home at night (ΔLog10 = 1.17 (0.38, 1.97)), and if any animal in the household was given antibiotics in the last 6 months (ΔLog10 = 1.16 (0.38, 1.93)) (Fig 2, Table S5a). Having an improved latrine, child antibiotic use, and child exposure to floors/soil were not associated with the abundance of ESBL-producing *E. coli* in child stool, while associations between the number of animals owned in the household or compound and the abundance of ESBL-producing *E. coli* in child stool were inconsistent (Fig 2, Tables S5a and 6).

**Figure 2.**
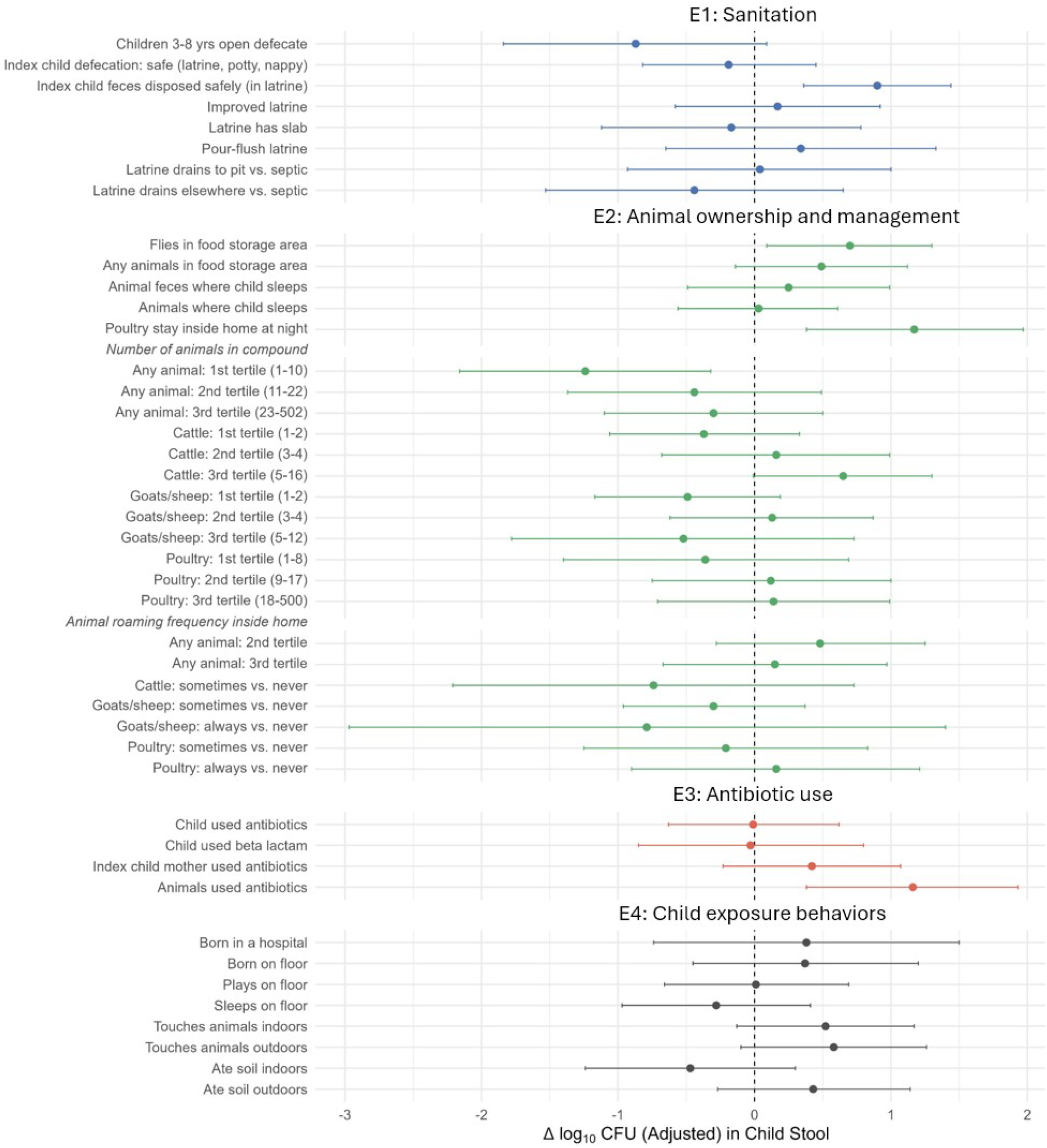
Associations between log10-transformed colony-forming units (CFU) of ESBL-producing *E. coli* in **child stool** vs. sanitation, animal ownership and management, antibiotic use, and child exposure behaviors. Adjusted analyses controlled for potential socio-demographic confounders (child age and sex, mother’s age and education, number of children <18 years in the household, number of individuals living in the compound, and asset-based wealth index) and study arm (intervention vs. control).

#### Chicken and cow feces

The abundance of ESBL-producing *E. coli* in chicken feces was higher in households where cows/buffalo roamed inside the home (ΔLog10 = 1.44 (0.12, 2.76)) and lower in households where children 3-8 years open defecated (ΔLog10 = −0.90 (−1.50, −0.29)) and households that had an improved latrine (ΔLog10 = −0.97 (−1.72, −0.22)) (Fig S1, Table S5b). The abundance of ESBL-producing *E. coli* in cow feces was not associated with sanitation characteristics or animal management practices (Fig S2, Table S5c). The abundance of ESBL-producing *E. coli* in chicken and cow feces increased with each additional 10 animals and each additional 10 chickens in the compound (Table S6).

#### Courtyard soil

The abundance of ESBL-producing *E. coli* in courtyard soil was 0.5-1 log higher in households where children 3-8 years open defecated (ΔLog10 = 1.22 (0.42, 2.03)), households where animal feces were observed on the floor of the child’s sleeping area (ΔLog10 = 0.69 (0.12, 1.25)) and households where animals roamed more often inside the home (ΔLog10 = 0.91 (0.18, 1.64)) (Fig S3, Table S5d). The abundance of ESBL-producing *E. coli* in courtyard soil 1.5-2 log lower in households that had a pour-flush latrine (ΔLog10 = −1.55 (−2.94, −0.16)) and a slab in the latrine (ΔLog10 = −1.95 (−3.44, −0.46)) (Fig S3, Table S5d). There were no associations between the number of animals in the household/compound and the abundance of ESBL-producing *E. coli* in courtyard soil (Table S6).

#### Floor swab

The abundance of ESBL-producing *E. coli* in floor swabs was approximately 1-log higher in households where cows/buffalo roamed inside the home (ΔLog10 = 1.06 (0.27, 1.85)), compounds in the highest tertile of cattle ownership (ΔLog10 = 0.66 (0.08, 1.24)) and households where animals were given antibiotics (ΔLog10 = 0.63 (0.02, 1.24)) (Fig S4, Table S5e). The abundance of ESBL-producing *E. coli* in floor swabs was lower in households where the latrine had a slab (ΔLog10 = −1.40 (−2.56, −0.25)) (Fig S4, Table S5e). The abundance of ESBL-producing *E. coli* in floor swabs increased with each additional cow in the household or compound (Table S6).

## Discussion

In our study in rural Bangladesh, almost all (96%) children aged 6 months were colonized with ESBL-producing *E. coli,* as well as almost all chickens and most cows. Our findings indicate a high burden of ESBL-producing *E. coli* colonization in our study setting and exceed prior estimates of 72-74% colonization prevalence within the first year of life among Bangladeshi children.^33,34^ Our study participants are from a low-income community in a riverine flood-prone area and have widespread animal ownership; these unique characteristics may explain the high AMR prevalence in this population. The colonization prevalence of 82-97% we observed among chickens and cows is consistent with other prior studies reporting high prevalence of ESBL-producing *E. coli* in livestock raised in small-scale settings in low- and middle-income countries.^20–24^ ESBL-producing *E. coli* is used by the WHO as an indicator to characterize the AMR burden across human, animal and environmental reservoirs;^43^ the high prevalence of colonization among children and domestic animals in our study signals a high burden of community-acquired AMR in this setting.

Among environmental samples, we detected ESBL-producing *E. coli* in approximately half of courtyard soil, third of indoor floors, <10% of child hands and stored food and 1% of drinking water samples. The highest prevalence among environmental samples being observed in soil and on floors corroborates growing evidence on the importance of these matrices as reservoirs of antimicrobial-resistant organisms and pathogens in low- and middle-income countries, especially in settings where backyard animal husbandry is common.^20,23^ Our previous studies in the same area in Bangladesh found cefotaxime-resistant *E. coli* and antimicrobial resistance genes in 89-100% of outdoor soil samples^17,42^ and 71% of indoor floor swabs,^18^ which is substantially higher than our findings. These previous studies were conducted during the monsoon season while the current study spanned a full year; seasonal trends may explain the difference in findings. While ESBL-producing *E. coli* has frequently been reported in ready-to-eat street foods, raw chicken, and fresh produce sold in markets,^12,44^ few studies have evaluated cooked food prepared and stored within households. One study in India found only one out of 12 cooked food samples to have ESBL-producing *E. coli*, similar to our findings.^12^ In our study, the prevalence of ESBL-producing *E. coli* in stored drinking water from tubewells was lower than previous studies in Bangladesh that detected ESBL-producing *E. coli* in tubewell water samples in a refugee camp (14%) and somewhat lower than previously observed among rural Bangladeshi households (5%).^45,46^ A prior study in Bangladesh found that among 46 households with infants aged <1 year, 11% (5 households) had overlapping antibiotic susceptibility patterns between child stool and drinking water samples, suggesting potential waterborne transmission of AMR;^35^ our findings indicate that drinking water exposure is unlikely to explain the high prevalence of colonization among children in our study setting.

Importantly, the abundance of ESBL-producing *E. coli* in child stool in our study was not significantly associated with the abundance or prevalence of ESBL-producing *E. coli* in any type of sample from the domestic environmental or animal feces, indicating colonization mechanisms not captured by the matrices sampled in our study, such as vertical transmission from colonized mothers or other environmentally mediated pathways.^35^ For example, among children <6 months, a prior study in rural Bangladesh found 60% of *E. coli* ingestion to be caused by mouthing contaminated objects.^31^ Given that the children in our study were very young (mean age: 0.6 years; range: 0.3–1.0 years), it is possible that transmission occurred through similar unmeasured pathways, such as contaminated objects frequently mouthed by infants, utensils/bottles used for feeding, complementary foods and liquids consumed by infants and mothers’ hands or nipples. However, this is unlikely given that the abundance of ESBL-producing *E. coli* in soil, on floors and on child hands did not appear to be associated with the abundance in child stool. A previous study in Peru suggested AMR transmission across children, animals, and domestic environments within households. In that study resistance profiles for similar antibiotics (tetracyclines) were shared between animal feces and water samples, and children’s drinking water showed resistance to the most commonly used antibiotic (ampicillin) used to treat children in the area.^47^ Domestic soils are particularly recognized as a dominant pathway for child exposure to pathogens and antimicrobial-resistant organisms.^17,48^ However, child exposure to soil measured in our study, such as the child being delivered on the floor, the frequency of child sleeping or playing on the floor and soil ingestion by the child, were not associated with the abundance of ESBL-producing *E. coli* in child stool.

Among household factors in the sanitation domain that we investigated as drivers of AMR, higher quality latrines (e.g. improved, pour-flush, with slabs) were generally associated with lower abundance of ESBL-producing *E. coli* across different matrices including chicken feces, courtyard soil and floor swabs. In a study of human fecal metagenomes from 26 countries, higher access to improved water and sanitation was associated with lower abundance of antimicrobial resistance genes in stool.^49^ In a recent study among peri-urban Malawian households, having a non-shared latrine was associated with lower abundance of cefotaxime-resistant *E. coli* in yard soil but the association was attenuated after controlling for household wealth.^50^ An intervention trial in Bangladesh found that children from households which received a combined water, sanitation, and handwashing intervention used less antibiotics, potentially indicating reduced AMR burden.^51^ However, the study did not measure AMR and did not isolate the effects of sanitation improvements from the effects of water quality and hygiene improvements. In contrast, child defecation and child feces management practices in our study had mixed associations with the abundance of ESBL-producing *E. coli* across matrices. Open defecation by children aged 3-8 years was associated with higher abundance in courtyard soil but lower abundance in chicken feces while disposal of index child feces in a latrine was associated with higher abundance in index child stool. Previous studies have also found mixed associations with unsafe child defecation and child feces disposal; these practices were associated with higher prevalence of *E. coli* on child hands, in stored water, and on both finished and unfinished floors^52,53^ but no association was found with diarrhea.^52^ Associations between AMR and young child defecation and management practices should be further investigated.

Among household factors in the animal ownership and management domain, animals entering the living space was generally associated with higher abundance of ESBL-producing *E. coli* across matrices; keeping chickens inside the home at night, cattle or chickens roaming inside the home and animals feces observed in the child’s sleeping area at the time of the household visit were associated with higher abundance in child stool, chicken feces, courtyard soil and floor swabs. These findings are consistent with growing evidence that domestic animals entering the indoor living space is a risk factor for the transmission of antimicrobial-resistant organisms and pathogens.^54,55^ In previous studies, animal health workers visiting the household and use of animal waste for gardening increased contamination in the household environment (doorknobs, soil, animal feeding equipment, water) with ESBL-producing *E. coli*.^19^ The number of animals in the household or compound was not consistently associated with the abundance of ESBL-producing *E. coli* in child feces in our study, while the abundance in cow/chicken feces and on floors increased with increasing number of chickens and cattle owned. These findings indicate that unsafe management of domestic animals, particularly access inside the home, may be a stronger driver of AMR colonization among children than the mere presence and number of animals. While the number of animals owned is a frequently used indicator to determine zoonotic risks from domestic animal husbandry, our findings support accumulating evidence that capturing specific animal practices is important to characterize infectious risks.^56,57^

Notably, the abundance of ESBL-producing *E. coli* in chicken feces, but not cow feces, was positively correlated with its abundance in courtyard soil and household floor swabs, suggesting that chicken feces may contribute more directly to contamination of the domestic environment or vice versa. However, higher number of cattle owned and cattle roaming inside the home were associated with higher abundance of ESBL-producing *E.* coli in child stool, chicken feces and floor swabs. Previous studies have identified chickens as a stronger contributor to domestic fecal contamination and adverse child health outcomes than other types of domestic animals.^20,22,23^ Chickens typically roam freely in household compounds to forage, and they deposit feces throughout shared living spaces, thus possibly increasing chances for environmental contamination from chicken feces, or the risk of chickens acquiring AMR from soils and floors. However, in our study, cattle appeared to be strong contributor to AMR. This could be due to our study area (Sirajganj) being the largest cattle-raising hub in Bangladesh. Finally, the abundance of ESBL-producing *E. coli* in courtyard soil and on floors was positively correlated with the abundance on child hands and in drinking water, similar with previous evidence that contamination from domestic soils and surfaces can permeate other domestic compartments.^25^

While 68% of index children in our study used antibiotics, and 26% used beta-lactam antibiotics in the last 6 months, child antibiotic use was not associated with the abundance of ESBL-producing *E. coli* in child stool or in any environmental matrix. In contrast, antibiotic use by domestic animals, reported by 22% of households that owned animals, was associated with higher abundance of ESBL-producing *E. coli* in child stool and on floors. Our findings are consistent with a prior study in rural Bangladesh that found that traditional clinical risk factors, such as recent antibiotic use or hospitalization, were not strongly associated with AMR colonization among infants.^33^ This also aligns with evidence from previous studies that humans and chickens share gut microbiomes,^28^ and antibiotic consumption in food-producing animals can contribute to the emergence and spread of AMR in human pathogens^27^.

Our findings should be interpreted with a few limitations in mind. Children in our study had very high prevalence of antimicrobial-resistant *E. coli* colonization. Therefore, we could not assess factors associated with the binary prevalence of colonization and only assessed associations with the abundance of ESBL-producing *E. coli* in stool. Most studies use the prevalence of colonization as the relevant endpoint; the clinical relevance of the abundance of resistant organisms per gram of stool is less clear, and factors driving the abundance of ESBL-producing *E. coli* in the gut may differ from factors that drive the binary probability of gut colonization. Our highly colonized population also limits the generalizability of our findings to settings with lower baseline levels of AMR. Further, a small sample size limited our ability to adjust for a larger number of potential confounders. Hence, residual confounding from unmeasured or imperfectly measured factors may have skewed our findings. Our small sample size, especially for samples that were not collected from all households due to lack of availability (food, chicken feces, cow feces), may also have limited our statistical power to detect associations. We did not collect samples from mothers, e.g. hand rinses, stool, vaginal flora, or other direct human-to-human transmission pathways within households. Therefore, we were unable to evaluate the relative contribution of transmission from caregivers or other household members to child colonization compared with environmental pathways. Additionally, we collected a one-time sample and were unable to assess colonization trends over time. Child stool samples were collected only one day after environmental samples; this temporal ordering is not sufficient to establish directionality of transmission (e.g. environmental contamination leading to child colonization vs. colonized children contaminating the environment). Studies with a larger sample size, more variable prevalence, and prospective longitudinal sampling may reveal clearer trends over time and as children age. Finally, we did not evaluate seasonal patterns. Bacterial persistence varies by rainfall, temperature, and other seasonal factors. However, our data collection spanned a year (May 2024–August 2025), thus capturing multiple seasons.

We found high prevalence of colonization with ESBL-producing *E. coli* among young children and domestic animals. The prevalence ESBL-producing *E. coli* was low along traditional fecal-oral transmission pathways, such as drinking water, food, and hands, but higher in domestic soils and on indoor floors, indicating that these matrices deserve attention as potential reservoirs of AMR. Among household characteristics, sanitation indicators and animal management practices were associated with levels of ESBL-producing *E.* coli across sampled matrices. Future studies should investigate whether sanitation improvements and interventions that limit free-roaming animals can reduce AMR in human, animal, and environmental reservoirs.

## Data Availability

All data produced in the present study are available upon reasonable request to the authors

## Acknowledgements

Research reported in this publication was supported by the National Institute of Child Health and Human Development under Award Number R01HD108196. The content is solely the responsibility of the authors and does not necessarily represent the official views of the National Institutes of Health. The study was also supported by a seed grant from the Global One Health Academy at North Carolina State University. JBC is a Chan Zuckerberg Biohub Investigator.

