## supporting information for "Antimicrobial-resistant *E. coli* in human, animal and environmental reservoirs in rural Bangladeshi households with young children"

**Table S1**. Lower limit of detection (LOD) across sample types

| Sample type | LOD (range)[a] | Reporting unit |
| --- | --- | --- |
| Drinking water | 1 | Per 100 mL |
| Food | 0.30 – 1.14 | Per 1 dry g |
| Child hands | 2.5 | Per 2 hands |
| Floor swabs | 30, 15 | Per 0.25 $m^2$ |
| Courtyard soil | 1.02 – 12.60 | Per 1 dry g |
| Chicken feces | 200 - 2000000 | Per 1 g |
| Cow feces | 200 | Per 1 g |
| Child stool | 400 - 40000000 | Per 1 g |

[a] Hyphenated values represent the minimum and maximum values for samples that have a range of LODs based on sample amount processed, dilution factor and moisture content.

**Table S2.** Exposure variables

| **Exposure domain 1: Sanitation** | Variable type | Values |
|---|---|---|
| Handling of human feces | | |
| Children <3 years open defecate | Excluded | 98% practiced open defecation |
| Children 3-8 years open defecate | Binary | 0: never, 1: sometimes/always |
| Children >9 years or adults open defecate | Excluded | 0% practiced open defecation |
| Index child defecates safely (in latrine, potty, nappy) | Binary | 0: uncontained, 1: potty, nappy, latrine |
| Index child feces disposed of safely (in latrine) | Binary | 0: unsafely disposed, 1: disposed in latrine |
| Improved latrine | Binary | 0: unimproved or no latrine, 1: improved |
| Latrine has slab | Binary | 0: no, 1: yes |
| Pour-flush latrine | Binary | 0: no, 1: yes |
| Latrine drains to | Categorical | 1: septic tank 2: pit latrine, 3: elsewhere |
| Human feces visible on floor where index child sleeps | Excluded | 99% had no feces |
| **Exposure domain 2: Animal ownership/management** | | |
| Any animals observed in food storage area | Binary | 0: no, 1: yes |
| Any animal feces observed in food storage area | Excluded | 98% had no animal feces in the area |
| Flies observed in food storage area | Binary | 0: no, 1: yes |
| Animal feces visible on floor where index child sleeps | Binary | 0: no, 1: yes |
| Any animals currently on floor where index child sleeps | Binary | 0: no, 1: yes |
| Chickens/ducks/pigeons stay inside home at night | Binary | 0: outside, 1: inside |
| Number of animals household owns | | |
| Any animals | Tertiles | 0: none, 1-3: tertiles |
| Cattle/buffalo | Tertiles | 0: none, 1-3: tertiles |
| Goats/sheep | Tertiles | 0: none, 1-3: tertiles |
| Chickens/ducks/pigeons | Tertiles | 0: none, 1-3: tertiles |
| Number of animals compound owns | | |
| Any animals | Tertiles | 0: none, 1-3: tertiles |
| Cattle/buffalo | Tertiles | 0: none, 1-3: tertiles |
| Goats/sheep | Tertiles | 0: none, 1-3: tertiles |
| Chickens/ducks/pigeons | Tertiles | 0: none, 1-3: tertiles |
| Animal roaming frequency inside home | | |
| Any animals | Tertiles | 1-3: tertiles |
| Cattle/buffalo | Categorical | 1: never, 2: sometimes, 3: always |
| Goats/sheep | Categorical | 1: never, 2: sometimes, 3: always |
| Chickens/ducks/pigeons | Categorical | 1: never, 2: sometimes, 3: always |
| Animal roaming frequency inside compound | | |
| Any animals | Tertiles | 1-3: tertiles |
| Cattle/buffalo | Categorical | 1: never, 2: sometimes, 3: always |
| Goats/sheep | Categorical | 1: never, 2: sometimes, 3: always |
| Chickens/ducks/pigeons | Categorical | 1: never, 2: sometimes, 3: always |
| **Exposure domain 3: Antibiotic use in last 6 months** | | |
| Index child mother used antibiotics | Binary | 0: no, 1: yes |
| Index child mother used beta lactam antibiotics | Excluded | 98% did not use |
| Index child used antibiotics | Binary | 0: no, 1: yes |
| Index child used beta lactam antibiotics | Binary | 0: no, 1: yes |
| Any animal in household used antibiotics | Binary | 0: no, 1: yes |
| **Exposure domain 4: Index child exposure behaviors** | | |
| Delivered in hospital | Binary | 0: no, 1: yes |
| Delivered on the floor | Binary | 0: no, 1: yes |
| How often child sleeps on the floor | Binary | 0: never, 1: every day/sometimes |
| How often child plays on the floor | Binary | 0: never, 1: every day/sometimes |
| How often child plays with/touches animals inside home | Binary | 0: never, 1: every day/sometimes |
| How often child plays with/touches animals outside home | Binary | 0: never, 1: every day/sometimes |
| Child has eaten any soil inside home today/yesterday | Binary | 1: yes on either day, 0: no on both days |
| Child has eaten any soil outside home today/yesterday | Binary | 1: yes on either day, 0: no on both days |

**Table S3.** Exposure domains included in models for each outcome domain

| Outcome domain | E1: Sanitation | E2: Animal ownership and management | E3: Antibiotic use | E4: Child exposure behaviors |
|---|---|---|---|---|
| O1: ESBL *E. coli* in environment | ✓ | ✓ | ✓ | |
| O2: ESBL *E. coli* in animals | ✓ | ✓ | ✓ | |
| O3: ESBL *E. coli* in children | ✓ | ✓ | ✓ | ✓ |

**Table S4.** Sanitation, animal ownership and management, antibiotic use, child exposure behaviors and demographic indicators among enrolled households (N=112)

| Exposure domain 1: Sanitation | N | % or mean | n or SD |
|---|---|---|---|
| Sanitation (handling of human feces) | | | |
| Improved latrine, % (n) | 112 | 85.7 | 96 |
| Human feces visible on floor where index child sleeps, % (n) | 112 | 0.9 | 1 |
| Frequency of open defecation by household members, % (n) | | | |
| Children <3 years | 112 | | |
| Never | | 0 | 0 |
| Occasionally | | 1.8 | 2 |
| Daily | | 98.2 | 110 |
| Children 3-8 years | 74 | | |
| Never | | 73 | 54 |
| Occasionally | | 6.8 | 5 |
| Daily | | 20.3 | 15 |
| Individuals >9 years | 43 | | |
| Never | | 100 | 43 |
| Occasionally | | 0 | 0 |
| Daily | | 0 | 0 |
| Index child defecates in safe location (potty, nappy, latrine), % (n) | 112 | 43.8 | 49 |
| Index child feces disposed of in latrine, % (n) | 111 | 23.4 | 26 |
| Exposure domain 2: Animal ownership and management | N | % or mean | n or SD |
| Animals observed in food storage area, % (n) | 112 | 8 | 9 |
| Animal feces observed in food storage area, % (n) | 112 | 1.8 | 2 |
| Flies observed in food storage area, % (n) | 103 | 35.9 | 37 |
| Animal feces visible on floor where index child sleeps, % (n) | 112 | 48.2 | 54 |
| Animals observed where index child sleeps, % (n) | 112 | 36.6 | 41 |
| Chickens stay inside the home at night, % (n) | 112 | 10.7 | 12 |
| Number of animals the compound owns, mean (SD) [a] | | | |
| Any animal | 112 | 20.1 | 48.4 |
| Cattle/buffalo | 112 | 1.9 | 2.6 |
| Goats/sheep | 112 | 1.4 | 2.3 |
| Chickens/ducks/pigeons | 112 | 16.8 | 48.1 |
| Percent of compounds that own, % (n) | | | |
| Any animal | 112 | 85.7 | 96 |
| Cattle/buffalo | 112 | 54.5 | 61 |
| Goats/sheep | 112 | 42.0 | 47 |
| Chickens/ducks/pigeons | 112 | 79.5 | 89 |
| Number of animals the household owns, mean (SD) | | | |
| Any animal | 112 | 9.9 | 11.6 |
| Cattle/buffalo | 112 | 1.4 | 2.4 |
| Goats/sheep | 112 | 0.9 | 1.6 |
| Chickens/ducks/pigeons | 112 | 7.7 | 10.7 |
| Percent of households that own, % (n) | | | |
| Any animal | 112 | 77.7 | 87 |
| Cattle/buffalo | 112 | 54.5 | 61 |
| Goats/sheep | 112 | 32.1 | 36 |
| Chickens/ducks/pigeons | 112 | 68.8 | 77 |
| Any animals roam free inside home, mean (SD) [b] | | | |
| 1st tertile | 36 | 3.8 | 0.4 |
| 2nd tertile | 43 | 5.0 | 0 |
| 3rd tertile | 33 | 6.3 | 0.5 |
| Any animals roam free in compound, mean (SD) [b] | | | |
| 1st tertile | 58 | 5.4 | 0.8 |
| 2nd tertile | 38 | 7.0 | 0 |
| 3rd tertile | 16 | 8.3 | 0.4 |

| Exposure domain 3: Antibiotic use | N | % or mean | n or SD |
|---|---|---|---|
| Antibiotics used in last 6 months, % (n) | | | |
| Index child | 112 | 67.9 | 76 |
| Index child mother | 112 | 7.1 | 8 |
| Animals | 96 | 22.9 | 22 |
| Beta-lactam antibiotics used in last 6 months, % (n) | | | |
| Index child | 112 | 25.9 | 29 |
| Index child mother | 112 | 1.8 | 2 |
| Exposure domain 4: Index child exposure behaviors | N | % or mean | n or SD |
| Delivered in hospital, % (n) | 112 | 7.1 | 8 |
| Delivered at home on the floor, % (n) | 112 | 48.2 | 54 |
| Ate any soil inside home today/yesterday, % (n) | 111 | 18.9 | 21 |
| Ate any soil outside home today/yesterday, % (n) | 111 | 35.1 | 39 |
| Sleeps on the floor, % (n) | 112 | | |
| Never | | 77.7 | 87 |
| Sometimes | | 7.1 | 8 |
| Everyday | | 15.2 | 17 |
| Plays on the floor, % (n) | 112 | | |
| Never | | 21.4 | 24 |
| Sometimes | | 16.1 | 18 |
| Everyday | | 62.5 | 70 |
| Plays with/touches animals inside home, % (n) | 112 | | |
| Never | | 89.3 | 100 |
| Sometimes | | 5.4 | 6 |
| Everyday | | 5.4 | 6 |
| Plays with/touches animals outside home, % (n) | 112 | | |
| Never | | 82.1 | 92 |
| Sometimes | | 9.8 | 11 |
| Everyday | | 8.0 | 9 |
| Demographics | N | % or mean | n or SD |
| Caregiver's age, mean (SD) | 112 | 24.3 | 5.9 |
| Caregiver's years of education, mean (SD) | 112 | 7 | 3.3 |
| Index child in years, mean (range) | 113 | 0.6 | 0.3 – 1.0 |
| Female index child, % (n) | 113 | 44.2 | 50 |
| Number of individuals ≤18 years living in household, median (range) | 112 | 1 | 0 – 7 |
| Number of total individuals living in compound, median (range) | 112 | 5 | 2 – 13 |
| Household characteristics, % (n) | 112 | | |
| Has electricity | | 110 | 98.2 |
| Has improved wall material [c] | | 110 | 98.2 |
| Household owns >1 of, % (n) | 112 | | |
| Television | | 13 | 11.6 |
| Bicycle | | 20 | 17.9 |
| Motorcycle | | 7 | 6.3 |
| Mobile phone | | 109 | 97.3 |
| Refrigerator | | 35 | 31.3 |

[a] Compound is a group of households around a central courtyard shared by extended families.

[b] Roaming frequency was scored as always (3 points), sometimes (2 points), and never (1 points) for different animal species. Points for cattle, goats/sheep, and poultry were summed to generate composite roaming scores. Tertiles, means and standard deviations were calculated from the composite scores.

[c] Improved walls included concrete and corrugated steel, and unimproved walls included soil/clay, leaves or thatch, and bamboo, based on local norms.

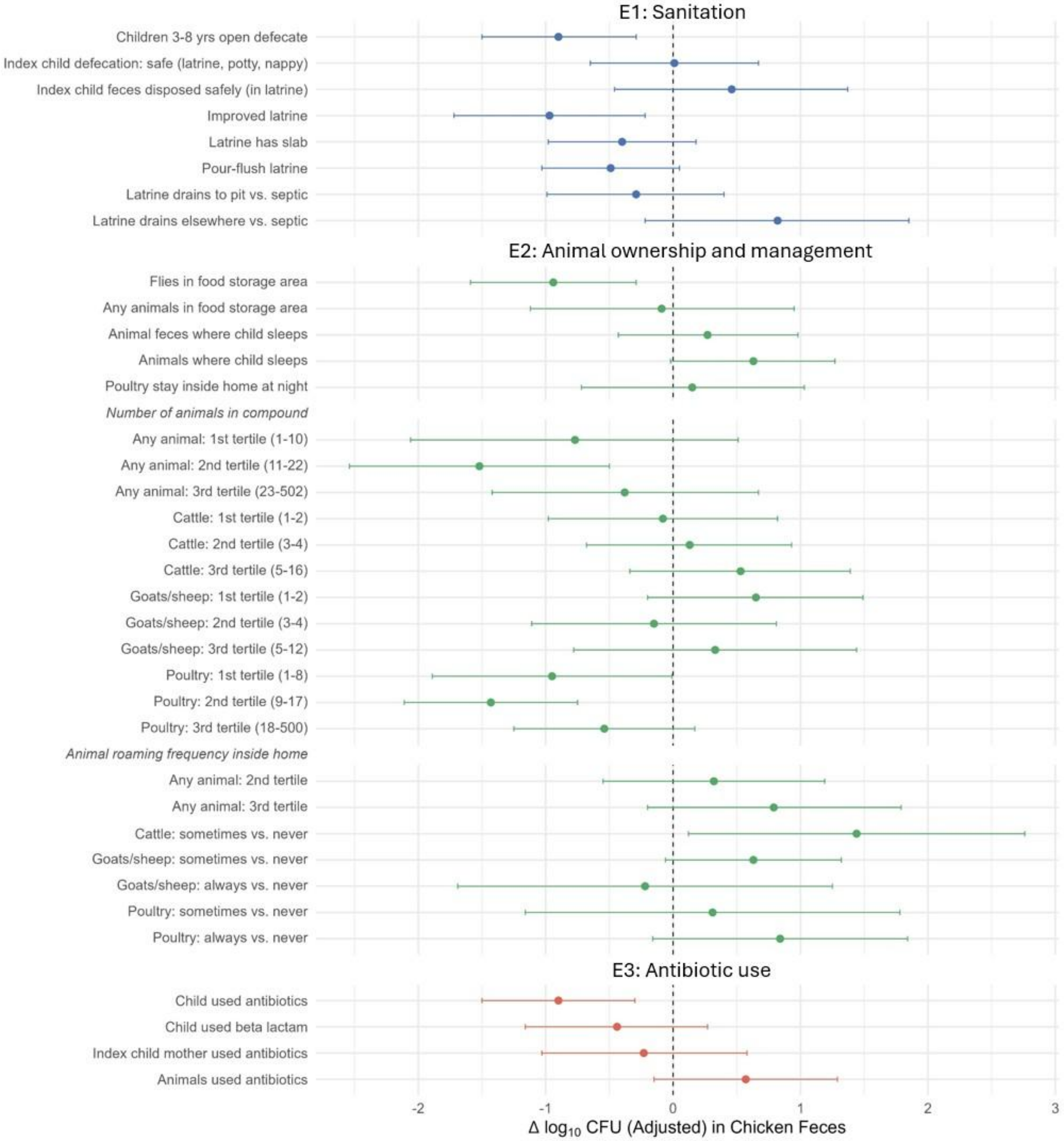

**Figure S1**. Associations between log10-transformed colony-forming units (CFU) of ESBL-producing *E. coli* in **chicken feces** vs. sanitation, animal ownership and management, and antibiotic use. Adjusted analyses controlled for potential socio-demographic confounders (mother's age and education, number of children <18 years in the household, number of individuals living in the compound, and asset-based wealth index) and study arm (intervention vs. control).

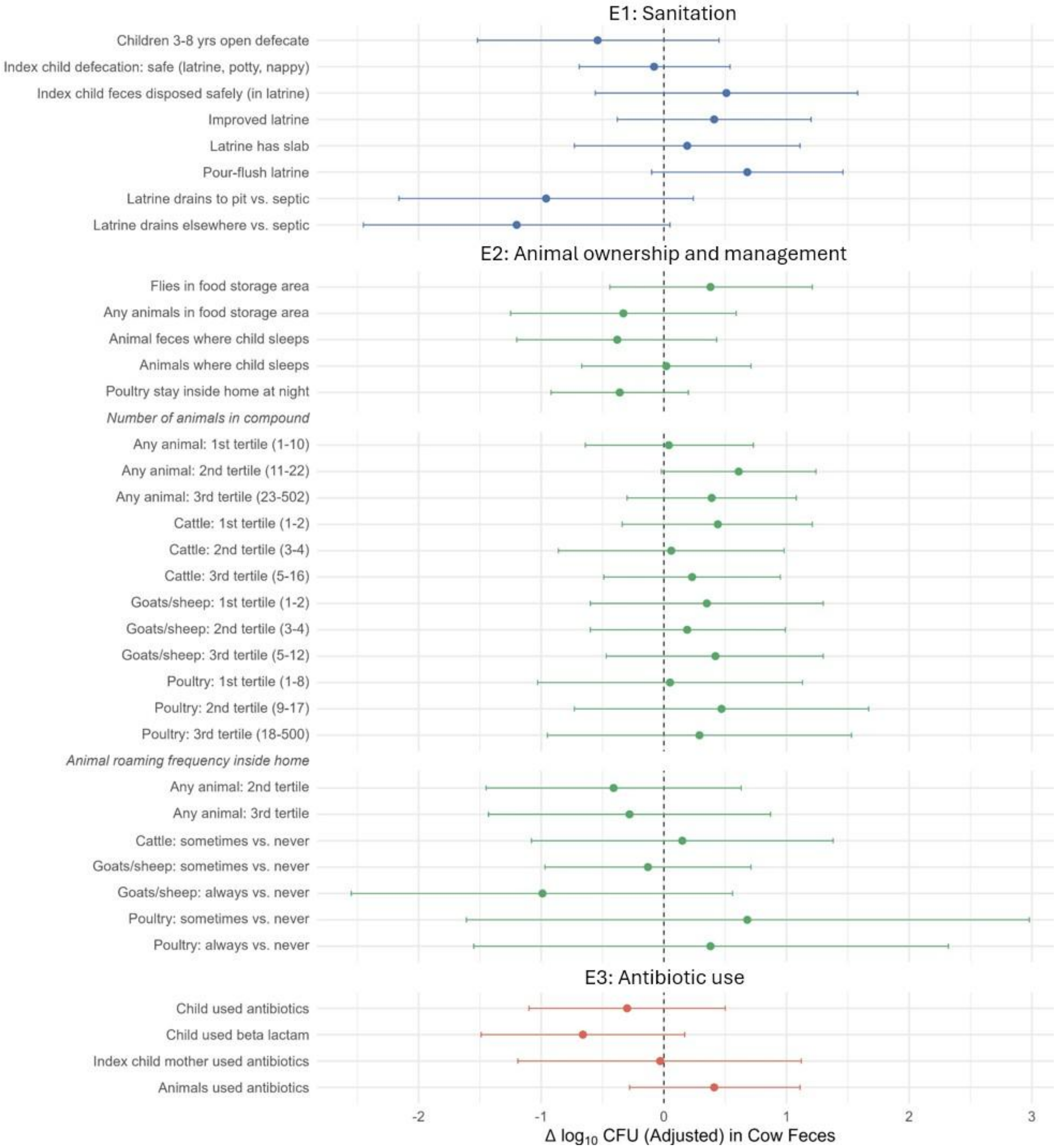

**Figure S2**. Associations between log10-transformed colony-forming units (CFU) of ESBL-producing *E. coli* in **cow feces** vs. sanitation, animal ownership and management, and antibiotic use. Adjusted analyses controlled for potential socio-demographic confounders (mother's age and education, number of children <18 years in the household, number of individuals living in the compound, and asset-based wealth index) and study arm (intervention vs. control).

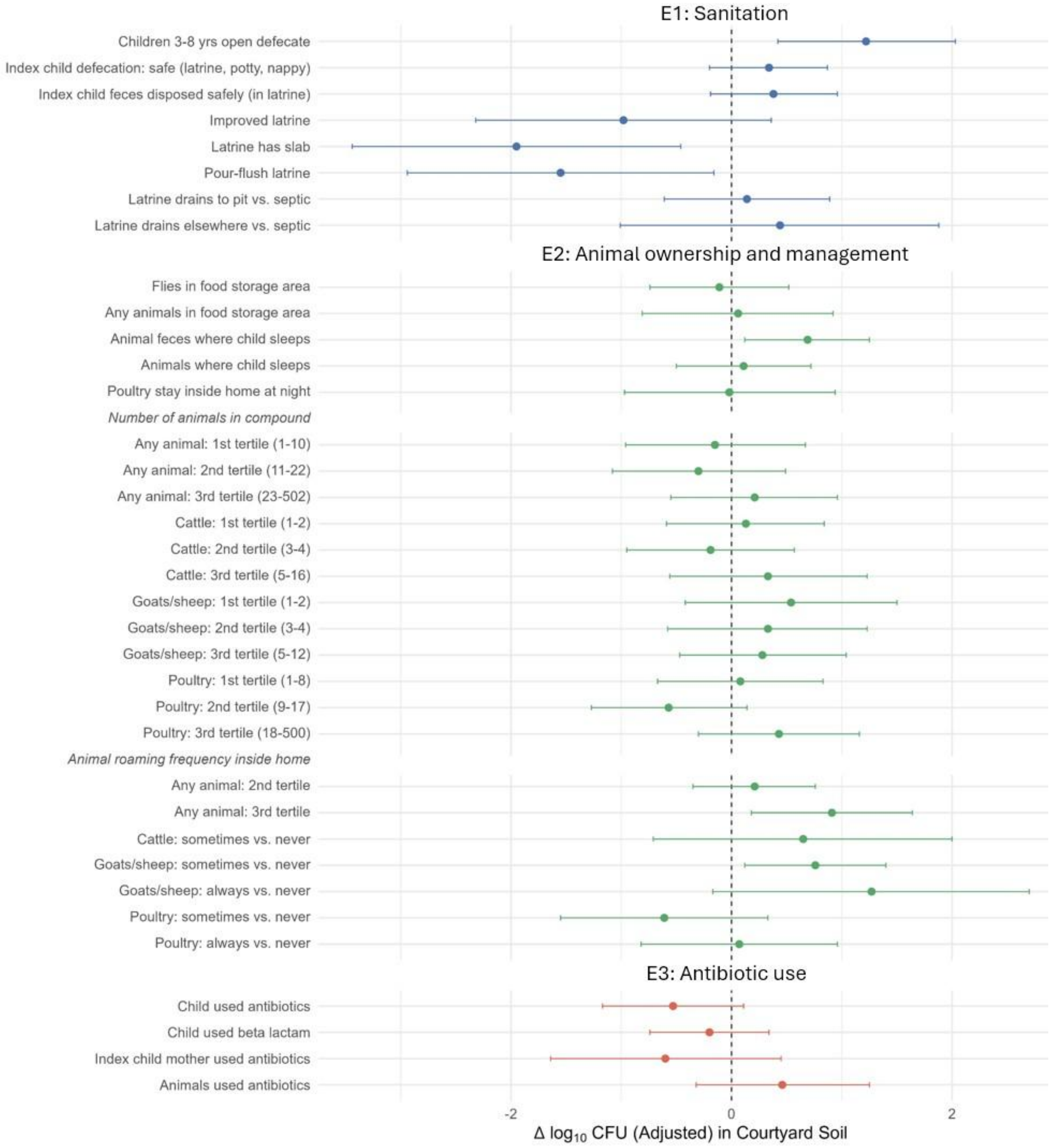

**Figure S3**. Associations between log10-transformed colony-forming units (CFU) of ESBL-producing *E. coli* in **courtyard soil** vs. sanitation, animal ownership and management, and antibiotic use. Adjusted analyses controlled for potential socio-demographic confounders (mother's age and education, number of children <18 years in the household, number of individuals living in the compound, and asset-based wealth index) and study arm (intervention vs. control).

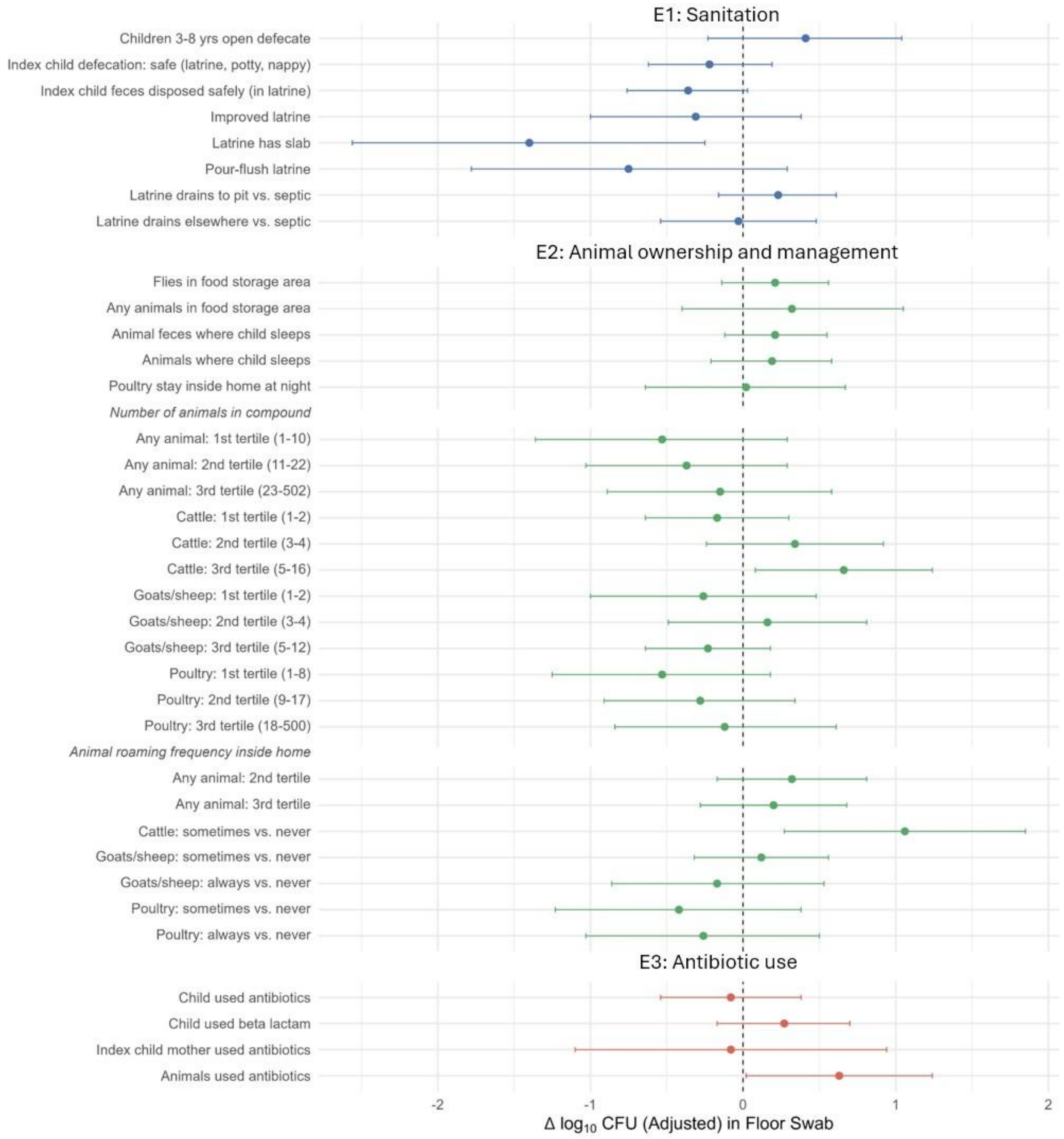

**Figure S4**. Associations between log10-transformed colony-forming units (CFU) of ESBL-producing *E. coli* in **floor swabs** vs. sanitation, animal ownership and management, and antibiotic use. Adjusted analyses controlled for potential socio-demographic confounders (mother's age and education, number of children <18 years in the household, number of individuals living in the compound, and asset-based wealth index) and study arm (intervention vs. control).

**Table S5a.** Associations between log10-transformed colony-forming units (CFU) of ESBL-producing *E. coli* in **child stool** vs. sanitation, animal ownership and management, antibiotic use, and child exposure behaviors. Adjusted analyses controlled for potential socio-demographic confounders (child age and sex, mother's age and education, number of children <18 years in the household, number of individuals living in the compound, and asset-based wealth index) and study arm (intervention vs. control).

| | | | | Unadjusted | | | Adjusted | | |
|---|---|---|---|---|---|---|---|---|---|
| | Exposure | N | Mean log10 CFU | ΔLog10 | 95% CI | p-value | ΔLog10 | 95% CI | p-value |
| **Exposure domain 1: Sanitation** | | | | | | | | | |
| Children 3–8 yrs open defecate | Never | 53 | 6.53 | ref | | | | | |
| | Sometimes/always | 20 | 5.86 | -0.67 | (-1.72, 0.37) | 0.21 | -0.87 | (-1.84, 0.09) | 0.08 |
| Index child defecation location | Unsafe | 63 | 6.60 | ref | | | | | |
| | Safe (latrine, nappy, potty) | 48 | 6.53 | -0.07 | (-0.68, 0.54) | 0.82 | -0.19 | (-0.82, 0.45) | 0.56 |
| Index child feces disposal | Unsafe | 83 | 6.36 | ref | | | | | |
| | Safely in latrine | 27 | 7.27 | **0.91** | **(0.46, 1.36)** | **<0.005** | **0.90** | **(0.36, 1.44)** | **<0.005** |
| Household has improved latrine | No | 15 | 6.34 | ref | | | | | |
| | Yes | 96 | 6.61 | 0.27 | (-0.55, 1.08) | 0.52 | 0.17 | (-0.58, 0.92) | 0.66 |
| Latrine has slab | No | 7 | 6.39 | ref | | | | | |
| | Yes | 104 | 6.58 | 0.19 | (-0.74, 1.12) | 0.69 | -0.17 | (-1.12, 0.78) | 0.73 |
| Pour-flush latrine | No | 8 | 6.09 | ref | | | | | |
| | Yes | 103 | 6.61 | 0.52 | (-0.42, 1.46) | 0.28 | 0.34 | (-0.65, 1.33) | 0.51 |
| Latrine drains to | Septic | 14 | 6.60 | ref | | | | | |
| | Pit latrine | 80 | 6.64 | 0.04 | (-1.00, 1.07) | 0.94 | 0.04 | (-0.93, 1.00) | 0.94 |
| | Elsewhere | 11 | 6.24 | -0.36 | (-1.66, 0.94) | 0.59 | -0.44 | (-1.53, 0.65) | 0.43 |
| **Exposure domain 2: Animal ownership and management** | | | | | | | | | |
| Flies in food storage area | Not observed | 64 | 6.30 | ref | | | | | |
| | Observed | 38 | 6.88 | **0.59** | **(-0.09, 1.26)** | **0.09** | **0.70** | **(0.09, 1.30)** | **0.02** |
| Any animals in food storage area | Not observed | 102 | 6.54 | | | | | | |
| | Observed | 9 | 6.97 | 0.43 | (-0.01, 0.88) | 0.06 | 0.49 | (-0.14, 1.12) | 0.13 |
| Animal feces on floor where index child sleeps | Not visible | 57 | 6.54 | ref | | | | | |
| | Visible | 54 | 6.61 | 0.07 | (-0.62, 0.76) | 0.85 | 0.25 | (-0.49, 0.99) | 0.51 |
| Any animals on floor where index child sleeps | Not present | 70 | 6.56 | ref | | | | | |
| | Present | 41 | 6.58 | 0.02 | (-0.64, 0.68) | 0.95 | 0.03 | (-0.56, 0.61) | 0.93 |
| Chickens/ducks/pigeons nighttime stay | Outside home | 99 | 6.47 | ref | | | | | |
| | Inside home | 12 | 7.39 | **0.92** | **(0.25, 1.59)** | **<0.005** | **1.17** | **(0.38, 1.97)** | **<0.005** |
| Number of animals household owns | | | | | | | | | |
| Any animals | None | 24 | 6.65 | ref | | | | | |
| | 1st tertile | 32 | 6.42 | -0.24 | (-1.17, 0.7) | 0.62 | -0.05 | (-0.92, 0.82) | 0.92 |
| | 2nd tertile | 30 | 6.7 | 0.05 | (-0.75, 0.84) | 0.91 | 0.08 | (-0.93, 1.08) | 0.88 |
| | 3rd tertile | 25 | 6.53 | -0.12 | (-0.91, 0.67) | 0.77 | -0.15 | (-1.04, 0.74) | 0.74 |
| Cattle/buffalo | None | 60 | 6.6 | ref | | | | | |
| | 1st tertile | 26 | 6.29 | -0.31 | (-1.1, 0.47) | 0.44 | -0.31 | (-0.99, 0.37) | 0.38 |
| | 2nd tertile | 11 | 7.14 | 0.54 | (-0.29, 1.36) | 0.20 | 0.54 | (-0.23, 1.31) | 0.17 |
| | 3rd tertile | 14 | 6.53 | -0.07 | (-1.1, 0.96) | 0.90 | -0.24 | (-1.31, 0.82) | 0.65 |
| Goats/sheep | None | 75 | 6.73 | ref | | | | | |
| | 1st tertile | 11 | 6.33 | -0.4 | (-1.65, 0.86) | 0.54 | -0.30 | (-1.35, 0.76) | 0.58 |
| | 2nd tertile | 14 | 6.32 | -0.41 | (-1.44, 0.62) | 0.43 | -0.72 | (-1.74, 0.30) | 0.16 |
| | 3rd tertile | 11 | 6.04 | -0.69 | (-1.9, 0.52) | 0.26 | -0.62 | (-1.67, 0.43) | 0.24 |

| | Exposure | N | Mean log10 CFU | Unadjusted | | | Adjusted | | |
|---|---|---|---|---|---|---|---|---|---|
| | | | | ΔLog10 | 95% CI | p-value | ΔLog10 | 95% CI | p-value |
| Chickens/ducks/pigeons | None | 33 | 6.4 | ref | | | | | |
| | 1st tertile | 26 | 6.87 | 0.47 | (-0.44, 1.38) | 0.31 | 0.63 | (-0.39, 1.65) | 0.23 |
| | 2nd tertile | 29 | 6.37 | -0.02 | (-0.79, 0.74) | 0.95 | -0.06 | (-0.87, 0.75) | 0.89 |
| | 3rd tertile | 23 | 6.73 | 0.34 | (-0.42, 1.09) | 0.38 | 0.36 | (-0.44, 1.17) | 0.38 |
| Number of animals compound owns | | | | | | | | | |
| Any animals | None | 16 | 7.01 | ref | | | | | |
| | 1st tertile | 33 | 6.03 | **-0.99** | **(-1.86, -0.11)** | **0.03** | **-1.24** | **(-2.16, -0.32)** | **0.008** |
| | 2nd tertile | 32 | 6.64 | -0.38 | (-1.15, 0.4) | 0.34 | -0.44 | (-1.37, 0.49) | 0.35 |
| | 3rd tertile | 30 | 6.86 | -0.15 | (-0.92, 0.62) | 0.70 | -0.30 | (-1.10, 0.50) | 0.46 |
| Cattle/buffalo | None | 50 | 6.52 | ref | | | | | |
| | 1st tertile | 24 | 6.1 | -0.42 | (-1.22, 0.38) | 0.30 | -0.37 | (-1.06, 0.33) | 0.30 |
| | 2nd tertile | 23 | 6.71 | 0.18 | (-0.72, 1.09) | 0.69 | 0.16 | (-0.68, 0.99) | 0.71 |
| | 3rd tertile | 14 | 7.33 | **0.81** | **(0.25, 1.37)** | **0.005** | **0.65** | **(-0.01, 1.30)** | **0.05** |
| Goats/sheep | None | 65 | 6.69 | ref | | | | | |
| | 1st tertile | 21 | 6.21 | -0.48 | (-1.41, 0.45) | 0.31 | -0.49 | (-1.17, 0.19) | 0.16 |
| | 2nd tertile | 15 | 6.87 | 0.18 | (-0.46, 0.81) | 0.59 | 0.13 | (-0.62, 0.87) | 0.73 |
| | 3rd tertile | 10 | 6.08 | -0.62 | (-1.95, 0.72) | 0.37 | -0.52 | (-1.78, 0.73) | 0.41 |
| Chickens/ducks/pigeons | None | 22 | 6.49 | ref | | | | | |
| | 1st tertile | 31 | 6.31 | -0.19 | (-1.2, 0.82) | 0.71 | -0.36 | (-1.40, 0.69) | 0.50 |
| | 2nd tertile | 31 | 6.79 | 0.29 | (-0.58, 1.16) | 0.51 | 0.12 | (-0.75, 1.00) | 0.78 |
| | 3rd tertile | 27 | 6.69 | 0.19 | (-0.71, 1.1) | 0.67 | 0.14 | (-0.71, 0.99) | 0.75 |
| Animal roaming frequency inside home | | | | | | | | | |
| Any animals | 1st tertile | 35 | 6.41 | ref | | | | | |
| | 2nd tertile | 44 | 6.82 | 0.41 | (-0.37, 1.19) | 0.30 | 0.48 | (-0.28, 1.25) | 0.21 |
| | 3rd tertile | 32 | 6.4 | -0.01 | (-0.85, 0.83) | 0.99 | 0.15 | (-0.67, 0.97) | 0.72 |
| Cattle/buffalo | Never | 104 | 6.61 | ref | | | | | |
| | Sometimes | 7 | 5.92 | -0.69 | (-2.29, 0.91) | 0.40 | -0.74 | (-2.21, 0.73) | 0.33 |
| Goats/sheep | Never | 76 | 6.72 | ref | | | | | |
| | Sometimes | 30 | 6.41 | -0.31 | (-1.04, 0.43) | 0.41 | -0.30 | (-0.96, 0.37) | 0.38 |
| | Always | 5 | 5.34 | -1.38 | (-3.06, 0.31) | 0.11 | -0.79 | (-2.97, 1.40) | 0.48 |
| Chickens/ducks/pigeons | Never | 8 | 6.54 | ref | | | | | |
| | Sometimes | 32 | 6.41 | -0.13 | (-1.49, 1.23) | 0.85 | -0.21 | (-1.25, 0.83) | 0.69 |
| | Always | 71 | 6.65 | 0.10 | (-1.23, 1.44) | 0.88 | 0.16 | (-0.90, 1.21) | 0.77 |
| Animal roaming frequency in compound | | | | | | | | | |
| Any animals | 1st tertile | 58 | 6.58 | ref | | | | | |
| | 2nd tertile | 37 | 6.73 | 0.15 | (-0.5, 0.81) | 0.65 | 0.45 | (-0.22, 1.13) | 0.19 |
| | 3rd tertile | 16 | 6.15 | -0.43 | (-1.15, 0.3) | 0.25 | -0.31 | (-1.03, 0.41) | 0.40 |
| Cattle/buffalo | Never | 69 | 6.62 | ref | | | | | |
| | Sometimes | 33 | 6.60 | -0.02 | (-0.75, 0.72) | 0.96 | 0.08 | (-0.64, 0.81) | 0.82 |
| | Always | 9 | 6.09 | -0.53 | (-1.77, 0.72) | 0.41 | -0.57 | (-1.73, 0.59) | 0.33 |
| Goats/sheep roam | Never | 3 | 4.89 | ref | | | | | |
| | Sometimes | 6 | 7.07 | 2.18 | (-0.11, 4.48) | 0.06 | 1.80 | (-0.22, 3.82) | 0.08 |
| | Always | 102 | 6.59 | 1.70 | (-0.55, 3.95) | 0.14 | 1.42 | (-0.43, 3.26) | 0.13 |
| Chickens/ducks/pigeons | Never | 29 | 6.63 | ref | | | | | |
| | Sometimes | 53 | 6.64 | 0.01 | (-0.77, 0.79) | 0.98 | 0.18 | (-0.72, 1.08) | 0.69 |
| | Always | 29 | 6.38 | -0.25 | (-1.05, 0.56) | 0.55 | 0.11 | (-0.59, 0.80) | 0.76 |
| **Exposure domain 3: Antibiotic use in last 6 months** | | | | | | | | | |
| Index child (any antibiotic) | Did not use | 35 | 6.49 | ref | | | | | |
| | Used | 76 | 6.61 | 0.12 | (-0.44, 0.69) | 0.68 | -0.01 | (-0.63, 0.62) | 0.98 |

| | Exposure | N | Mean log10 CFU | Unadjusted | | | Adjusted | | |
|---|---|---|---|---|---|---|---|---|---|
| | | | | ΔLog10 | 95% CI | p-value | ΔLog10 | 95% CI | p-value |
| Index child (beta-lactam) | Did not use | 82 | 6.56 | ref | | | | | |
| | Used | 29 | 6.61 | 0.05 | (-0.70, 0.81) | 0.89 | -0.03 | (-0.85, 0.80) | 0.95 |
| Index mother (any antibiotic) | Did not use | 103 | 6.57 | ref | | | | | |
| | Used | 8 | 6.59 | 0.02 | (-0.73, 0.77) | 0.97 | 0.42 | (-0.23, 1.07) | 0.20 |
| Any animal in household | Did not use | 74 | 6.31 | ref | | | | | |
| | Used | 21 | 7.15 | **0.84** | **(0.09, 1.58)** | **0.03** | **1.16** | **(0.38, 1.93)** | **<0.005** |
| **Exposure domain 4: Index child exposure behaviors** | | | | | | | | | |
| Delivery in hospital | Yes | 103 | 6.53 | ref | | | | | |
| | No | 8 | 7.15 | 0.62 | (-0.47, 1.71) | 0.26 | 0.38 | (-0.74, 1.50) | 0.51 |
| Delivery location | Elsewhere | 59 | 6.57 | ref | | | | | |
| | On the floor | 52 | 6.57 | -0.00 | (-0.65, 0.65) | 0.99 | 0.37 | (-0.45, 1.20) | 0.38 |
| Plays on the floor | Never | 24 | 6.56 | ref | | | | | |
| | Sometimes/always | 87 | 6.58 | 0.02 | (-0.61, 0.65) | 0.95 | 0.01 | (-0.66, 0.69) | 0.97 |
| Sleeps on floor | Never | 86 | 6.64 | ref | | | | | |
| | Sometimes/always | 25 | 6.32 | -0.32 | (-1.08, 0.43) | 0.40 | -0.28 | (-0.97, 0.41) | 0.43 |
| Plays with/touches animals inside home | No | 99 | 6.52 | ref | | | | | |
| | Yes | 12 | 6.98 | 0.46 | (-0.06, 0.98) | 0.08 | 0.52 | (-0.13, 1.17) | 0.12 |
| Plays with/touches animals outside home | No | 91 | 6.47 | ref | | | | | |
| | Yes | 20 | 7.04 | 0.57 | (-0.10, 1.25) | 0.10 | 0.58 | (-0.10, 1.26) | 0.09 |
| Ate any soil inside home in last 2 days | Has not eaten | 90 | 6.59 | ref | | | | | |
| | Has eaten | 20 | 6.44 | -0.15 | (-0.81, 0.51) | 0.66 | -0.47 | (-1.24, 0.30) | 0.23 |
| Ate any soil outside home in last 2 days | Has not eaten | 70 | 6.40 | ref | | | | | |
| | Has eaten | 40 | 6.84 | 0.43 | (-0.15, 1.02) | 0.14 | 0.43 | (-0.27, 1.14) | 0.23 |

**Table S5b**. Associations between log10-transformed colony-forming units (CFU) of ESBL-producing *E. coli* in **chicken feces** vs. sanitation, animal ownership and management, and antibiotic use. Adjusted analyses controlled for potential socio-demographic confounders (mother's age and education, number of children <18 years in the household, number of individuals living in the compound, and asset-based wealth index) and study arm (intervention vs. control).

| | | | | Unadjusted | | | Adjusted | | |
|---|---|---|---|---|---|---|---|---|---|
| | Exposure | N | Mean log10 CFU | ΔLog10 | 95% CI | p-value | ΔLog10 | 95% CI | p-value |
| **Exposure domain 1: Sanitation** | | | | | | | | | |
| Children 3–8 yrs open defecate | Never | 33 | 5.90 | ref | | | | | |
| | Sometimes/always | 14 | 5.09 | **-0.81** | **(-1.56, -0.07)** | **0.03** | **-0.90** | **(-1.50, -0.29)** | **<0.005** |
| Index child defecation location | Unsafe | 39 | 5.46 | ref | | | | | |
| | Safe (latrine, nappy, potty) | 26 | 5.37 | -0.08 | (-0.75, 0.58) | 0.80 | 0.01 | (-0.65, 0.67) | 0.98 |
| Index child feces disposal | Unsafe | 50 | 5.39 | ref | | | | | |
| | Safely in latrine | 14 | 5.58 | 0.19 | (-0.58, 0.96) | 0.63 | 0.46 | (-0.46, 1.37) | 0.33 |
| Household has improved latrine | No | 9 | 6.28 | ref | | | | | |
| | Yes | 56 | 5.29 | **-1.00** | **(-1.57, -0.42)** | **<0.005** | **-0.97** | **(-1.72, -0.22)** | **0.01** |
| Latrine has slab | No | 6 | 5.99 | ref | | | | | |
| | Yes | 59 | 5.37 | **-0.63** | **(-1.15, -0.11)** | **0.02** | -0.40 | (-0.98, 0.18) | 0.17 |
| Pour-flush latrine | No | 7 | 5.94 | ref | | | | | |
| | Yes | 58 | 5.36 | **-0.58** | **(-1.08, -0.08)** | **0.02** | -0.49 | (-1.03, 0.05) | 0.08 |
| Latrine drains to | Septic | 10 | 5.48 | ref | | | | | |
| | Pit latrine | 44 | 5.23 | -0.25 | (-1.07, 0.57) | 0.55 | -0.29 | (-0.99, 0.40) | 0.41 |
| | Elsewhere | 6 | 6.17 | 0.69 | (-0.31, 1.69) | 0.18 | 0.82 | (-0.22, 1.85) | 0.12 |
| **Exposure domain 2: Animal ownership and management** | | | | | | | | | |
| Flies in food storage area | Not observed | 38 | 5.64 | ref | | | | | |
| | Observed | 21 | 5.25 | -0.39 | (-1.01, 0.23) | 0.22 | **-0.94** | **(-1.59, -0.29)** | **0.01** |
| Any animals in food storage area | Not observed | 58 | 5.45 | ref | | | | | |
| | Observed | 7 | 5.23 | -0.21 | (-1.26, 0.84) | 0.69 | -0.09 | (-1.12, 0.95) | 0.87 |
| Animal feces on floor where index child sleeps | Not visible | 25 | 5.19 | ref | | | | | |
| | Visible | 40 | 5.57 | 0.37 | (-0.27, 1.01) | 0.26 | 0.27 | (-0.43, 0.98) | 0.45 |
| Any animals on floor where index child sleeps | Not present | 32 | 5.21 | ref | | | | | |
| | Present | 33 | 5.63 | 0.42 | (-0.26, 1.10) | 0.23 | 0.63 | (-0.02, 1.27) | 0.06 |
| Chickens/ducks/pigeons nighttime stay | Outside home | 56 | 5.42 | ref | | | | | |
| | Inside home | 9 | 5.47 | 0.05 | (-0.76, 0.86) | 0.90 | 0.15 | (-0.72, 1.03) | 0.73 |
| Number of animals household owns | | | | | | | | | |
| Any animals | None | 5 | 5.35 | ref | | | | | |
| | 1st tertile | 15 | 5.61 | 0.26 | (-0.97, 1.49) | 0.68 | -0.08 | (-1.40, 1.24) | 0.91 |
| | 2nd tertile | 24 | 5.47 | 0.12 | (-1.14, 1.38) | 0.85 | -0.03 | (-1.40, 1.33) | 0.96 |
| | 3rd tertile | 21 | 5.26 | -0.09 | (-1.43, 1.25) | 0.89 | -0.19 | (-1.48, 1.10) | 0.77 |
| Cattle/buffalo | None | 26 | 5.46 | ref | | | | | |
| | 1st tertile | 17 | 5.37 | -0.09 | (-0.95, 0.78) | 0.84 | 0.11 | (-0.75, 0.98) | 0.80 |
| | 2nd tertile | 11 | 5.51 | 0.05 | (-0.82, 0.92) | 0.91 | 0.10 | (-0.89, 1.09) | 0.84 |
| | 3rd tertile | 11 | 5.33 | -0.14 | (-1.12, 0.85) | 0.79 | -0.11 | (-1.03, 0.81) | 0.81 |
| Goats/sheep | None | 39 | 5.31 | ref | | | | | |
| | 1st tertile | 8 | 6.19 | **0.88** | **(0.08, 1.68)** | **0.03** | 0.68 | (-0.33, 1.68) | 0.19 |
| | 2nd tertile | 8 | 5.79 | 0.48 | (-0.35, 1.3) | 0.26 | 0.36 | (-0.72, 1.44) | 0.51 |
| | 3rd tertile | 10 | 4.94 | -0.37 | (-1.24, 0.49) | 0.40 | -0.54 | (-1.47, 0.39) | 0.26 |
| Chickens/ducks/pigeons | None | 7 | 5.27 | ref | | | | | |

| | Exposure | N | Mean log10 CFU | Unadjusted | | | Adjusted | | |
|---|---|---|---|---|---|---|---|---|---|
| | | | | ΔLog10 | 95% CI | p-value | ΔLog10 | 95% CI | p-value |
| | 1st tertile | 16 | 5.42 | 0.15 | (-0.95, 1.26) | 0.79 | -0.12 | (-1.29, 1.04) | 0.83 |
| | 2nd tertile | 23 | 5.71 | 0.43 | (-0.75, 1.62) | 0.47 | 0.50 | (-0.61, 1.61) | 0.38 |
| | 3rd tertile | 19 | 5.14 | -0.14 | (-1.39, 1.12) | 0.83 | -0.17 | (-1.33, 0.99) | 0.77 |
| Number of animals compound owns | | | | | | | | | |
| Any animals | None | 1 | 5.86 | ref | | | | | |
| | 1st tertile | 16 | 5.62 | -0.24 | (-0.82, 0.34) | 0.42 | -0.77 | (-2.06, 0.51) | 0.24 |
| | 2nd tertile | 27 | 4.98 | **-0.87** | **(-1.38, -0.37)** | **<0.005** | **-1.52** | **(-2.54, -0.50)** | **<0.005** |
| | 3rd tertile | 21 | 5.82 | -0.04 | (-0.55, 0.48) | 0.89 | -0.38 | (-1.42, 0.67) | 0.48 |
| Cattle/buffalo | None | 21 | 5.38 | ref | | | | | |
| | 1st tertile | 15 | 5.12 | -0.26 | (-1.21, 0.69) | 0.60 | -0.08 | (-0.98, 0.82) | 0.86 |
| | 2nd tertile | 19 | 5.44 | 0.07 | (-0.74, 0.88) | 0.87 | 0.13 | (-0.68, 0.93) | 0.76 |
| | 3rd tertile | 10 | 5.94 | 0.56 | (-0.37, 1.5) | 0.24 | 0.53 | (-0.34, 1.39) | 0.24 |
| Goats/sheep | None | 32 | 5.16 | ref | | | | | |
| | 1st tertile | 14 | 6.01 | **0.84** | **(0.07, 1.62)** | **0.03** | 0.65 | (-0.20, 1.49) | 0.13 |
| | 2nd tertile | 11 | 5.23 | 0.07 | (-0.7, 0.84) | 0.86 | -0.15 | (-1.11, 0.81) | 0.76 |
| | 3rd tertile | 8 | 5.69 | 0.53 | (-0.44, 1.49) | 0.29 | 0.33 | (-0.78, 1.44) | 0.56 |
| Chickens/ducks/pigeons | None | 2 | 6.23 | ref | | | | | |
| | 1st tertile | 19 | 5.51 | -0.72 | (-1.52, 0.09) | 0.08 | -0.95 | (-1.89, -0.01) | 0.05 |
| | 2nd tertile | 24 | 4.98 | **-1.25** | **(-1.99, -0.51)** | **<0.005** | **-1.43** | **(-2.11, -0.75)** | **<0.005** |
| | 3rd tertile | 20 | 5.79 | -0.44 | (-1.17, 0.29) | 0.24 | -0.54 | (-1.25, 0.17) | 0.14 |
| Animal roaming frequency inside home | | | | | | | | | |
| Any animals | 1st tertile | 15 | 5.06 | | | | | | |
| | 2nd tertile | 29 | 5.3 | 0.24 | (-0.58, 1.06) | 0.57 | 0.32 | (-0.55, 1.19) | 0.47 |
| | 3rd tertile | 21 | 5.85 | 0.79 | (-0.01, 1.6) | 0.05 | 0.79 | (-0.20, 1.79) | 0.12 |
| Cattle/buffalo | Never | 62 | 5.37 | ref | | | | | |
| | Sometimes | 3 | 6.62 | **1.25** | **(0.35, 2.15)** | **0.006** | **1.44** | **(0.12, 2.76)** | **0.03** |
| Goats/sheep | Never | 43 | 5.22 | ref | | | | | |
| | Sometimes | 18 | 5.84 | 0.62 | (-0.02, 1.26) | 0.06 | 0.63 | (-0.06, 1.32) | 0.07 |
| | Always | 4 | 5.76 | 0.54 | (-0.21, 1.29) | 0.16 | -0.22 | (-1.69, 1.25) | 0.77 |
| Chickens/ducks/pigeons | Never | 1 | 4.93 | ref | | | | | |
| | Sometimes | 16 | 5.11 | 0.18 | (-0.46, 0.81) | 0.58 | 0.31 | (-1.16, 1.78) | 0.68 |
| | Always | 48 | 5.54 | **0.61** | **(0.23, 1.00)** | **<0.005** | 0.84 | (-0.16, 1.84) | 0.10 |
| Animal roaming frequency in compound | | | | | | | | | |
| Any animals | 1st tertile | 32 | 5.16 | ref | | | | | |
| | 2nd tertile | 24 | 5.67 | 0.51 | (-0.12, 1.14) | 0.11 | 0.57 | (-0.19, 1.34) | 0.14 |
| | 3rd tertile | 9 | 5.71 | 0.56 | (-0.59, 1.71) | 0.34 | 0.58 | (-0.53, 1.68) | 0.31 |
| Cattle/buffalo | Never | 41 | 5.31 | ref | | | | | |
| | Sometimes | 21 | 5.74 | 0.43 | (-0.17, 1.03) | 0.16 | 0.33 | (-0.32, 0.99) | 0.32 |
| | Always | 3 | 4.71 | -0.60 | (-3.07, 1.86) | 0.63 | -0.52 | (-2.69, 1.65) | 0.64 |
| Goats/sheep roam | Sometimes | 1 | 5.86 | ref | | | | | |
| | Always | 64 | 5.42 | **-0.44** | **(-0.79, -0.09)** | **0.01** | -0.87 | (-1.84, 0.11) | 0.08 |
| Chickens/ducks/pigeons | Never | 15 | 5.02 | ref | | | | | |
| | Sometimes | 31 | 5.52 | 0.50 | (-0.37, 1.37) | 0.26 | 0.31 | (-0.62, 1.24) | 0.51 |
| | Always | 19 | 5.59 | 0.58 | (-0.38, 1.54) | 0.24 | 0.50 | (-0.67, 1.67) | 0.40 |
| **Exposure domain 3: Antibiotic use in last 6 months** | | | | | | | | | |
| Index child (any antibiotic) | Did not use | 23 | 5.95 | ref | | | | | |
| | Used | 42 | 5.14 | -0.81 | (-1.48, -0.15) | 0.02 | **-0.90** | **(-1.50, -0.30)** | **<0.005** |
| Index child (beta-lactam) | Did not use | 52 | 5.50 | ref | | | | | |
| | Used | 13 | 5.11 | -0.47 | (-1.09, 0.15) | 0.14 | -0.44 | (-1.16, 0.27) | 0.22 |

| | Exposure | N | Mean log10 CFU | Unadjusted | | | Adjusted | | |
|---|---|---|---|---|---|---|---|---|---|
| | | | | ΔLog10 | 95% CI | p-value | ΔLog10 | 95% CI | p-value |
| Index mother (any antibiotic) | Did not use | 61 | 5.43 | ref | | | | | |
| | Used | 4 | 5.38 | -0.04 | (-0.75, 0.66) | 0.91 | -0.23 | (-1.03, 0.58) | 0.58 |
| Any animal in household | Did not use | 46 | 5.24 | ref | | | | | |
| | Used | 18 | 5.86 | -0.39 | (-1.15, 0.36) | 0.31 | 0.57 | (-0.15, 1.29) | 0.12 |

**Table S5c**. Associations between log10-transformed colony-forming units (CFU) of ESBL-producing *E. coli* in **cow feces** vs. sanitation, animal ownership and management, and antibiotic use. Adjusted analyses controlled for potential socio-demographic confounders (mother's age and education, number of children <18 years in the household, number of individuals living in the compound, and asset-based wealth index) and study arm (intervention vs. control).

| | | | | Unadjusted | | | Adjusted | | |
|---|---|---|---|---|---|---|---|---|---|
| | Exposure | N | Mean log10 CFU | ΔLog10 | 95% CI | p-value | ΔLog10 | 95% CI | p-value |
| **Exposure domain 1: Sanitation** | | | | | | | | | |
| Children 3–8 yrs open defecate | Never | 30 | 3.65 | ref | | | | | |
| | Sometimes/always | 15 | 3.36 | -0.29 | (-1.17, 0.59) | 0.51 | -0.54 | (-1.52, 0.45) | 0.28 |
| Index child defecation location | Unsafe | 34 | 3.56 | ref | | | | | |
| | Safe (latrine, nappy, potty) | 28 | 3.66 | 0.09 | (-0.65, 0.83) | 0.81 | -0.08 | (-0.69, 0.54) | 0.80 |
| Index child feces disposal | Unsafe | 49 | 3.56 | ref | | | | | |
| | Safely in latrine | 13 | 3.79 | 0.23 | (-0.72, 1.18) | 0.64 | 0.51 | (-0.56, 1.58) | 0.35 |
| Household has improved latrine | No | 8 | 3.20 | ref | | | | | |
| | Yes | 54 | 3.67 | 0.47 | (-0.43, 1.37) | 0.31 | 0.41 | (-0.38, 1.20) | 0.31 |
| Latrine has slab | No | 3 | 3.38 | ref | | | | | |
| | Yes | 59 | 3.62 | 0.24 | (-0.78, 1.25) | 0.65 | 0.19 | (-0.73, 1.11) | 0.69 |
| Pour-flush latrine | No | 3 | 2.63 | ref | | | | | |
| | Yes | 59 | 3.66 | **1.03** | **(0.12, 1.93)** | **0.03** | 0.68 | (-0.10, 1.46) | 0.09 |
| Latrine drains to | Septic | 8 | 4.37 | ref | | | | | |
| | Pit latrine | 45 | 3.58 | -0.80 | (-1.75, 0.16) | 0.10 | -0.96 | (-2.16, 0.24) | 0.12 |
| | Elsewhere | 7 | 3.10 | **-1.27** | **(-2.47, -0.07)** | **0.04** | -1.20 | (-2.45, 0.05) | 0.06 |
| **Exposure domain 2: Animal ownership and management** | | | | | | | | | |
| Flies in food storage area | Not observed | 35 | 3.50 | ref | | | | | |
| | Observed | 21 | 3.66 | 0.16 | (-0.64, 0.96) | 0.70 | 0.38 | (-0.44, 1.21) | 0.36 |
| Any animals in food storage area | Not observed | 56 | 3.61 | ref | | | | | |
| | Observed | 6 | 3.61 | 0.01 | (-1.28, 1.3) | 0.99 | -0.33 | (-1.25, 0.59) | 0.48 |
| Animal feces on floor where index child sleeps | Not visible | 22 | 3.74 | ref | | | | | |
| | Visible | 40 | 3.53 | -0.20 | (-1.00, 0.60) | 0.62 | -0.38 | (-1.20, 0.43) | 0.35 |
| Any animals on floor where index child sleeps | Not present | 30 | 3.49 | ref | | | | | |
| | Present | 32 | 3.71 | 0.22 | (-0.52, 0.96) | 0.56 | 0.02 | (-0.67, 0.71) | 0.95 |
| Chickens/ducks/pigeons nighttime stay | Outside home | 53 | 3.67 | ref | | | | | |
| | Inside home | 9 | 3.23 | -0.43 | (-1.16, 0.29) | 0.24 | -0.36 | (-0.92, 0.20) | 0.21 |
| Number of animals household owns | | | | | | | | | |
| Any animals | None | 7 | 3.97 | ref | | | | | |
| | 1st tertile | 17 | 3.32 | -0.65 | (-1.98, 0.68) | 0.34 | -1.09 | (-2.38, 0.21) | 0.10 |
| | 2nd tertile | 20 | 3.34 | -0.63 | (-1.91, 0.65) | 0.33 | **-1.40** | **(-2.77, -0.04)** | **0.04** |
| | 3rd tertile | 18 | 4.03 | 0.06 | (-1.51, 1.62) | 0.94 | -0.64 | (-1.92, 0.65) | 0.33 |
| Cattle/buffalo | None | 11 | 3.71 | ref | | | | | |
| | 1st tertile | 25 | 3.6 | -0.12 | (-1.13, 0.9) | 0.82 | -0.64 | (-1.74, 0.46) | 0.26 |
| | 2nd tertile | 12 | 3.48 | -0.24 | (-1.3, 0.83) | 0.66 | -1.09 | (-2.38, 0.20) | 0.10 |
| | 3rd tertile | 14 | 3.64 | -0.07 | (-1.4, 1.25) | 0.91 | -0.89 | (-2.03, 0.25) | 0.12 |
| Goats/sheep | None | 14 | 3.72 | ref | | | | | |
| | 1st tertile | 15 | 3.43 | -0.29 | (-1.49, 0.92) | 0.64 | 0.46 | (-0.72, 1.64) | 0.44 |
| | 2nd tertile | 20 | 3.63 | -0.09 | (-1.14, 0.97) | 0.87 | 0.32 | (-0.94, 1.58) | 0.62 |
| | 3rd tertile | 13 | 3.64 | -0.07 | (-1.46, 1.31) | 0.92 | -0.31 | (-1.33, 0.71) | 0.55 |
| Chickens/ducks/pigeons | None | 36 | 3.57 | ref | | | | | |
| | 1st tertile | 9 | 3.86 | 0.29 | (-0.88, 1.46) | 0.63 | -0.62 | (-1.87, 0.62) | 0.32 |

| | Exposure | N | Mean log10 CFU | Unadjusted | | | Adjusted | | |
|---|---|---|---|---|---|---|---|---|---|
| | | | | ΔLog10 | 95% CI | p-value | ΔLog10 | 95% CI | p-value |
| | 2nd tertile | 9 | 3.77 | 0.2 | (-0.78, 1.18) | 0.69 | -0.53 | (-1.61, 0.54) | 0.33 |
| | 3rd tertile | 8 | 3.29 | -0.29 | (-1.35, 0.78) | 0.60 | -0.66 | (-1.92, 0.59) | 0.30 |
| Number of animals compound owns | | | | | | | | | |
| Any animals | None | 2 | 3.23 | ref | | | | | |
| | 1st tertile | 18 | 3.32 | 0.09 | (-0.45, 0.63) | 0.75 | 0.04 | (-0.64, 0.73) | 0.90 |
| | 2nd tertile | 22 | 4.02 | **0.80** | **(0.19, 1.41)** | **0.01** | 0.61 | (-0.02, 1.24) | 0.06 |
| | 3rd tertile | 20 | 3.45 | 0.22 | (-0.33, 0.77) | 0.44 | 0.39 | (-0.30, 1.08) | 0.26 |
| Cattle/buffalo | None | 3 | 3 | ref | | | | | |
| | 1st tertile | 22 | 3.79 | **0.78** | **(0.05, 1.52)** | **0.04** | 0.44 | (-0.34, 1.21) | 0.27 |
| | 2nd tertile | 23 | 3.58 | 0.58 | (-0.05, 1.21) | 0.07 | 0.06 | (-0.86, 0.98) | 0.90 |
| | 3rd tertile | 14 | 3.49 | 0.48 | (-0.26, 1.22) | 0.20 | 0.23 | (-0.49, 0.95) | 0.53 |
| Goats/sheep | None | 30 | 3.55 | ref | | | | | |
| | 1st tertile | 14 | 3.73 | 0.18 | (-0.73, 1.09) | 0.70 | 0.35 | (-0.60, 1.30) | 0.47 |
| | 2nd tertile | 10 | 3.59 | 0.04 | (-0.78, 0.86) | 0.92 | 0.19 | (-0.60, 0.99) | 0.64 |
| | 3rd tertile | 8 | 3.64 | 0.09 | (-0.91, 1.1) | 0.85 | 0.42 | (-0.47, 1.30) | 0.35 |
| Chickens/ducks/pigeons | None | 7 | 3.38 | ref | | | | | |
| | 1st tertile | 17 | 3.5 | 0.12 | (-0.97, 1.21) | 0.83 | 0.05 | (-1.03, 1.13) | 0.92 |
| | 2nd tertile | 22 | 3.84 | 0.46 | (-0.71, 1.62) | 0.44 | 0.47 | (-0.73, 1.67) | 0.44 |
| | 3rd tertile | 16 | 3.49 | 0.11 | (-1.22, 1.44) | 0.87 | 0.29 | (-0.95, 1.53) | 0.64 |
| Animal roaming frequency inside home | | | | | | | | | |
| Any animals | 1st tertile | 13 | 4.05 | ref | | | | | |
| | 2nd tertile | 30 | 3.47 | -0.58 | (-1.65, 0.49) | 0.29 | -0.41 | (-1.45, 0.63) | 0.44 |
| | 3rd tertile | 19 | 3.52 | -0.53 | (-1.74, 0.67) | 0.39 | -0.28 | (-1.43, 0.87) | 0.63 |
| Cattle/buffalo | Never | 58 | 3.60 | ref | | | | | |
| | Sometimes | 4 | 3.64 | 0.04 | (-1.09, 1.16) | 0.95 | 0.15 | (-1.08, 1.38) | 0.81 |
| Goats/sheep | Never | 42 | 3.69 | ref | | | | | |
| | Sometimes | 18 | 3.46 | -0.23 | (-1.10, 0.64) | 0.60 | -0.13 | (-0.97, 0.71) | 0.76 |
| | Always | 2 | 3.14 | -0.55 | (-2.27, 1.17) | 0.53 | -0.99 | (-2.55, 0.56) | 0.21 |
| Chickens/ducks/pigeons | Never | 4 | 3.55 | ref | | | | | |
| | Sometimes | 11 | 3.98 | 0.43 | (-1.69, 2.55) | 0.69 | 0.68 | (-1.61, 2.98) | 0.56 |
| | Always | 47 | 3.52 | -0.02 | (-2.00, 1.95) | 0.98 | 0.38 | (-1.55, 2.32) | 0.70 |
| Animal roaming frequency in compound | | | | | | | | | |
| Any animals | 1st tertile | 26 | 3.78 | ref | | | | | |
| | 2nd tertile | 24 | 3.2 | -0.58 | (-1.33, 0.17) | 0.13 | -0.54 | (-1.31, 0.24) | 0.17 |
| | 3rd tertile | 12 | 4.06 | 0.29 | (-0.72, 1.3) | 0.58 | 0.41 | (-0.55, 1.36) | 0.40 |
| Cattle/buffalo | Never | 29 | 3.50 | ref | | | | | |
| | Sometimes | 24 | 3.69 | 0.18 | (-0.56, 0.92) | 0.63 | 0.40 | (-0.25, 1.04) | 0.23 |
| | Always | 9 | 3.71 | 0.21 | (-0.84, 1.26) | 0.70 | 0.10 | (-1.03, 1.23) | 0.86 |
| Goats/sheep roam | Never | 2 | 4.08 | ref | | | | | |
| | Sometimes | 2 | 3.23 | -0.85 | (-4.15, 2.45) | 0.61 | -0.75 | (-3.93, 2.42) | 0.64 |
| | Always | 58 | 3.60 | -0.48 | (-3.84, 2.88) | 0.78 | -0.34 | (-3.53, 2.85) | 0.83 |
| Chickens/ducks/pigeons | Never | 15 | 4.32 | ref | | | | | |
| | Sometimes | 31 | 3.43 | **-0.89** | **(-1.68, -0.11)** | **0.03** | -0.41 | (-1.16, 0.34) | 0.28 |
| | Always | 16 | 3.27 | **-1.06** | **(-1.99, -0.12)** | **0.03** | -0.87 | (-1.85, 0.11) | 0.08 |
| **Exposure domain 3: Antibiotic use in last 6 months** | | | | | | | | | |
| Index child (any antibiotic) | Did not use | 19 | 3.58 | ref | | | | | |
| | Used | 43 | 3.62 | 0.04 | (-0.64, 0.72) | 0.91 | -0.30 | (-1.10, 0.50) | 0.47 |
| Index child (beta-lactam) | Did not use | 46 | 3.73 | ref | | | | | |
| | Used | 16 | 3.26 | -0.47 | (-1.09, 0.15) | 0.14 | -0.66 | (-1.49, 0.17) | 0.12 |

| | Exposure | N | Mean log10 CFU | Unadjusted | | | Adjusted | | |
|---|---|---|---|---|---|---|---|---|---|
| | | | | ΔLog10 | 95% CI | p-value | ΔLog10 | 95% CI | p-value |
| Index mother (any antibiotic) | Did not use | 57 | 3.63 | ref | | | | | |
| | Used | 5 | 3.32 | -0.32 | (-1.31, 0.68) | 0.54 | -0.03 | (-1.19, 1.12) | 0.96 |
| Any animal in household | Did not use | 40 | 3.49 | ref | | | | | |
| | Used | 20 | 3.87 | 0.38 | (-0.23, 0.99) | 0.22 | 0.41 | (-0.28, 1.11) | 0.25 |

**Table S5d**. Associations between log10-transformed colony-forming units (CFU) of ESBL-producing *E. coli* in **courtyard soil** vs. sanitation, animal ownership and management, and antibiotic use. Adjusted analyses controlled for potential socio-demographic confounders (mother's age and education, number of children <18 years in the household, number of individuals living in the compound, and asset-based wealth index) and study arm (intervention vs. control).

| | | | | Unadjusted | | | Adjusted | | |
|---|---|---|---|---|---|---|---|---|---|
| | Exposure | N | Mean log10 CFU | ΔLog10 | 95% CI | p-value | ΔLog10 | 95% CI | p-value |
| **Exposure domain 1: Sanitation** | | | | | | | | | |
| Children 3–8 yrs open defecate | Never | 54 | 0.69 | ref | | | | | |
| | Sometimes/always | 20 | 2.05 | **1.36** | **(0.31, 2.41)** | **0.01** | **1.22** | **(0.42, 2.03)** | **<0.005** |
| Index child defecation location | Unsafe | 63 | 0.95 | ref | | | | | |
| | Safe (latrine, nappy, potty) | 49 | 1.15 | 0.20 | (-0.43, 0.83) | 0.53 | 0.34 | (-0.20, 0.87) | 0.22 |
| Index child feces disposal | Unsafe | 85 | 0.97 | ref | | | | | |
| | Safely in latrine | 26 | 1.22 | 0.25 | (-0.43, 0.92) | 0.48 | 0.38 | (-0.19, 0.96) | 0.19 |
| Household has improved latrine | No | 15 | 1.94 | ref | | | | | |
| | Yes | 97 | 0.90 | -1.05 | (-2.53, 0.43) | 0.17 | -0.98 | (-2.32, 0.36) | 0.15 |
| Latrine has slab | No | 7 | 2.73 | ref | | | | | |
| | Yes | 105 | 0.92 | **-1.81** | **(-3.50, -0.12)** | **0.04** | **-1.95** | **(-3.44, -0.46)** | **0.01** |
| Pour-flush latrine | No | 8 | 2.30 | ref | | | | | |
| | Yes | 104 | 0.94 | -1.36 | (-2.93, 0.21) | 0.09 | **-1.55** | **(-2.94, -0.16)** | **0.03** |
| Latrine drains to | Septic | 14 | 0.88 | ref | | | | | |
| | Pit latrine | 81 | 0.90 | 0.02 | (-0.69, 0.74) | 0.95 | 0.14 | (-0.61, 0.89) | 0.71 |
| | Elsewhere | 11 | 1.26 | 0.38 | (-1.29, 2.05) | 0.65 | 0.44 | (-1.01, 1.88) | 0.55 |
| **Exposure domain 2: Animal ownership and management** | | | | | | | | | |
| Flies in food storage area | Not observed | 66 | 0.96 | ref | | | | | |
| | Observed | 37 | 1.15 | 0.19 | (-0.46, 0.83) | 0.57 | -0.11 | (-0.74, 0.52) | 0.74 |
| Any animals in food storage area | Not observed | 103 | 1.05 | ref | | | | | |
| | Observed | 9 | 0.82 | -0.23 | (-1.1, 0.63) | 0.59 | 0.06 | (-0.81, 0.92) | 0.90 |
| Animal feces on floor where index child sleeps | Not visible | 58 | 0.69 | ref | | | | | |
| | Visible | 54 | 1.41 | **0.73** | **(0.15, 1.31)** | **0.01** | **0.69** | **(0.12, 1.25)** | **0.02** |
| Any animals on floor where index child sleeps | Not present | 71 | 1.06 | ref | | | | | |
| | Present | 41 | 0.99 | -0.08 | (-0.69, 0.54) | 0.81 | 0.11 | (-0.50, 0.72) | 0.72 |
| Chickens/ducks/pigeons nighttime stay | Outside home | 100 | 1.07 | ref | | | | | |
| | Inside home | 12 | 0.73 | -0.34 | (-1.41, 0.73) | 0.53 | -0.02 | (-0.97, 0.94) | 0.97 |
| Number of animals household owns | | | | | | | | | |
| Any animals | None | 25 | 0.76 | | | | | | |
| | 1st tertile | 33 | 0.89 | 0.12 | (-0.6, 0.85) | 0.74 | -0.33 | (-0.99, 0.34) | 0.33 |
| | 2nd tertile | 29 | 1.48 | 0.71 | (-0.09, 1.51) | 0.08 | 0.51 | (-0.35, 1.38) | 0.24 |
| | 3rd tertile | 25 | 1 | 0.23 | (-0.43, 0.9) | 0.49 | 0.14 | (-0.60, 0.88) | 0.71 |
| Cattle/buffalo | None | 61 | 0.8 | | | | | | |
| | 1st tertile | 25 | 1.43 | 0.63 | (-0.16, 1.42) | 0.12 | 0.50 | (-0.28, 1.28) | 0.21 |
| | 2nd tertile | 12 | 1.56 | 0.76 | (-0.31, 1.82) | 0.16 | 0.45 | (-0.64, 1.53) | 0.42 |
| | 3rd tertile | 14 | 0.93 | 0.13 | (-0.69, 0.95) | 0.76 | -0.21 | (-1.02, 0.60) | 0.61 |
| Goats/sheep | None | 76 | 0.79 | | | | | | |
| | 1st tertile | 11 | 1.71 | 0.92 | (-0.53, 2.38) | 0.21 | 0.40 | (-0.84, 1.63) | 0.53 |
| | 2nd tertile | 14 | 1.81 | **1.02** | **(0.16, 1.88)** | **0.02** | 0.62 | (-0.25, 1.48) | 0.16 |
| | 3rd tertile | 11 | 1.08 | 0.29 | (-0.64, 1.23) | 0.54 | 0.17 | (-0.78, 1.13) | 0.72 |
| Chickens/ducks/pigeons | None | 35 | 0.9 | | | | | | |

| | Exposure | N | Mean log10 CFU | Unadjusted | | | Adjusted | | |
|---|---|---|---|---|---|---|---|---|---|
| | | | | ΔLog10 | 95% CI | p-value | ΔLog10 | 95% CI | p-value |
| | 1st tertile | 26 | 1.19 | 0.29 | (-0.6, 1.18) | 0.52 | 0.07 | (-0.73, 0.86) | 0.87 |
| | 2nd tertile | 29 | 0.92 | 0.02 | (-0.61, 0.66) | 0.94 | 0.16 | (-0.51, 0.83) | 0.64 |
| | 3rd tertile | 22 | 1.22 | 0.32 | (-0.39, 1.02) | 0.38 | 0.30 | (-0.50, 1.10) | 0.46 |
| Number of animals compound owns | | | | | | | | | |
| Any animals | None | 16 | 0.71 | | | | | | |
| | 1st tertile | 35 | 1.2 | 0.48 | (-0.34, 1.31) | 0.25 | -0.15 | (-0.96, 0.67) | 0.72 |
| | 2nd tertile | 31 | 0.82 | 0.11 | (-0.7, 0.92) | 0.79 | -0.30 | (-1.08, 0.49) | 0.46 |
| | 3rd tertile | 30 | 1.24 | 0.53 | (-0.17, 1.24) | 0.14 | 0.21 | (-0.55, 0.96) | 0.59 |
| Cattle/buffalo | None | 51 | 0.89 | | | | | | |
| | 1st tertile | 23 | 1.18 | 0.29 | (-0.45, 1.02) | 0.44 | 0.13 | (-0.59, 0.84) | 0.73 |
| | 2nd tertile | 24 | 0.99 | 0.11 | (-0.66, 0.88) | 0.79 | -0.19 | (-0.95, 0.57) | 0.63 |
| | 3rd tertile | 14 | 1.42 | 0.53 | (-0.53, 1.59) | 0.33 | 0.33 | (-0.56, 1.23) | 0.46 |
| Goats/sheep | None | 65 | 0.74 | | | | | | |
| | 1st tertile | 21 | 1.73 | **0.98** | **(0.07, 1.9)** | **0.04** | 0.54 | (-0.42, 1.50) | 0.27 |
| | 2nd tertile | 15 | 1.36 | 0.62 | (-0.37, 1.6) | 0.22 | 0.33 | (-0.58, 1.23) | 0.48 |
| | 3rd tertile | 11 | 1.01 | 0.27 | (-0.52, 1.06) | 0.50 | 0.28 | (-0.47, 1.04) | 0.46 |
| Chickens/ducks/pigeons | None | 23 | 0.89 | | | | | | |
| | 1st tertile | 32 | 1.34 | 0.44 | (-0.38, 1.26) | 0.29 | 0.08 | (-0.67, 0.83) | 0.83 |
| | 2nd tertile | 30 | 0.46 | -0.44 | (-1.16, 0.29) | 0.24 | -0.57 | (-1.27, 0.14) | 0.12 |
| | 3rd tertile | 27 | 1.44 | 0.55 | (-0.21, 1.31) | 0.16 | 0.43 | (-0.30, 1.16) | 0.25 |
| Animal roaming frequency inside home | | | | | | | | | |
| Any animals | 1st tertile | 36 | 0.62 | | | | | | |
| | 2nd tertile | 43 | 0.85 | 0.23 | (-0.27, 0.74) | 0.36 | 0.21 | (-0.35, 0.76) | 0.47 |
| | 3rd tertile | 33 | 1.73 | **1.11** | **(0.3, 1.91)** | **0.01** | **0.91** | **(0.18, 1.64)** | **0.01** |
| Cattle/buffalo | Never | 105 | 1.01 | ref | | | | | |
| | Sometimes | 7 | 1.46 | 0.46 | (-1.12, 2.03) | 0.57 | 0.65 | (-0.71, 2.00) | 0.35 |
| Goats/sheep | Never | 76 | 0.72 | ref | | | | | |
| | Sometimes | 31 | 1.69 | **0.97** | **(0.23, 1.71)** | **0.01** | **0.76** | **(0.12, 1.40)** | **0.02** |
| | Always | 5 | 1.79 | 1.07 | (-0.46, 2.60) | 0.17 | 1.27 | (-0.17, 2.70) | 0.08 |
| Chickens/ducks/pigeons | Never | 9 | 1.22 | ref | | | | | |
| | Sometimes | 32 | 0.41 | -0.81 | (-1.78, 0.16) | 0.10 | -0.61 | (-1.55, 0.33) | 0.21 |
| | Always | 71 | 1.30 | 0.08 | (-0.87, 1.03) | 0.87 | 0.07 | (-0.82, 0.96) | 0.88 |
| Animal roaming frequency in compound | | | | | | | | | |
| Any animals | 1st tertile | 58 | 1.04 | | | | | | |
| | 2nd tertile | 38 | 0.84 | -0.2 | (-0.81, 0.41) | 0.52 | -0.35 | (-0.99, 0.28) | 0.28 |
| | 3rd tertile | 16 | 1.5 | 0.46 | (-0.49, 1.42) | 0.34 | 0.26 | (-0.70, 1.22) | 0.60 |
| Cattle/buffalo | Never | 69 | 1.01 | ref | | | | | |
| | Sometimes | 33 | 1.14 | 0.13 | (-0.51, 0.77) | 0.69 | -0.07 | (-0.76, 0.61) | 0.83 |
| | Always | 10 | 0.86 | -0.15 | (-1.03, 0.74) | 0.74 | -0.34 | (-1.14, 0.46) | 0.41 |
| Goats/sheep roam | Never | 3 | 1.53 | ref | | | | | |
| | Sometimes | 6 | 0.92 | -0.61 | (-2.23, 1.01) | 0.46 | -0.16 | (-1.61, 1.28) | 0.83 |
| | Always | 103 | 1.03 | -0.50 | (-2.00, 0.99) | 0.51 | -0.55 | (-1.82, 0.71) | 0.39 |
| Chickens/ducks/pigeons | Never | 30 | 0.90 | ref | | | | | |
| | Sometimes | 53 | 1.04 | 0.14 | (-0.56, 0.84) | 0.69 | -0.18 | (-0.81, 0.46) | 0.59 |
| | Always | 29 | 1.17 | 0.27 | (-0.52, 1.06) | 0.50 | 0.21 | (-0.62, 1.04) | 0.61 |
| **Exposure domain 3: Antibiotic use in last 6 months** | | | | | | | | | |
| Index child (any antibiotic) | Did not use | 36 | 1.51 | ref | | | | | |
| | Used | 76 | 0.81 | **-0.69** | **(-1.34, -0.04)** | **0.04** | -0.53 | (-1.17, 0.11) | 0.10 |
| Index child (beta-lactam) | Did not use | 83 | 1.10 | ref | | | | | |

| | Exposure | N | Mean log10 CFU | Unadjusted | | | Adjusted | | |
|---|---|---|---|---|---|---|---|---|---|
| | | | | ΔLog10 | 95% CI | p-value | ΔLog10 | 95% CI | p-value |
| | Used | 29 | 0.86 | -0.24 | (-0.85, 0.38) | 0.45 | -0.20 | (-0.74, 0.34) | 0.46 |
| Index mother (any antibiotic) | Did not use | 104 | 1.08 | | | | | | |
| | Used | 8 | 0.48 | -0.59 | (-1.5, 0.31) | 0.20 | -0.60 | (-1.64, 0.45) | 0.26 |
| Any animal in household | Did not use | 74 | 0.89 | ref | | | | | |
| | Used | 22 | 1.75 | **0.86** | **(0.10, 1.62)** | **0.03** | 0.46 | (-0.32, 1.25) | 0.24 |

**Table S5e**. Associations between log10-transformed colony-forming units (CFU) of ESBL-producing *E. coli* in **floor swabs** vs. sanitation, animal ownership and management, and antibiotic use. Adjusted analyses controlled for potential socio-demographic confounders (mother's age and education, number of children <18 years in the household, number of individuals living in the compound, and asset-based wealth index) and study arm (intervention vs. control).

| | | | | Unadjusted | | | Adjusted | | |
|---|---|---|---|---|---|---|---|---|---|
| | Exposure | N | Mean log10 CFU | ΔLog10 | 95% CI | p-value | ΔLog10 | 95% CI | p-value |
| **Exposure domain 1: Sanitation** | | | | | | | | | |
| Children 3–8 yrs open defecate | Never | 54 | 1.81 | ref | | | | | |
| | Sometimes/always | 20 | 2.20 | 0.40 | (-0.30, 1.10) | 0.26 | 0.41 | (-0.23, 1.04) | 0.21 |
| Index child defecation location | Unsafe | 63 | 1.84 | ref | | | | | |
| | Safe (latrine, nappy, potty) | 49 | 1.47 | -0.37 | (-0.82, 0.08) | 0.11 | -0.22 | (-0.62, 0.19) | 0.30 |
| Index child feces disposal | Unsafe | 85 | 1.82 | ref | | | | | |
| | Safely in latrine | 26 | 1.25 | **-0.57** | **(-0.98, -0.17)** | **0.006** | -0.36 | (-0.76, 0.03) | 0.07 |
| Household has improved latrine | No | 15 | 1.99 | ref | | | | | |
| | Yes | 97 | 1.63 | -0.36 | (-1.16, 0.44) | 0.38 | -0.31 | (-1.00, 0.38) | 0.38 |
| Latrine has slab | No | 7 | 3.07 | ref | | | | | |
| | Yes | 105 | 1.59 | **-1.48** | **(-2.77, -0.18)** | **0.02** | **-1.40** | **(-2.56, -0.25)** | **0.02** |
| Pour-flush latrine | No | 8 | 2.48 | ref | | | | | |
| | Yes | 104 | 1.62 | -0.86 | (-2.02, 0.30) | 0.14 | -0.75 | (-1.78, 0.29) | 0.16 |
| Latrine drains to | Septic | 14 | 1.48 | ref | | | | | |
| | Pit latrine | 81 | 1.67 | 0.19 | (-0.23, 0.61) | 0.37 | 0.23 | (-0.16, 0.61) | 0.25 |
| | Elsewhere | 11 | 1.41 | -0.06 | (-0.57, 0.44) | 0.80 | -0.03 | (-0.54, 0.48) | 0.91 |
| **Exposure domain 2: Animal ownership and management** | | | | | | | | | |
| Flies in food storage area | Not observed | 66 | 1.46 | ref | | | | | |
| | Observed | 37 | 1.99 | **0.52** | **(0.13, 0.92)** | **0.01** | 0.21 | (-0.14, 0.56) | 0.24 |
| Any animals in food storage area | Not observed | 103 | 1.68 | | | | | | |
| | Observed | 9 | 1.75 | 0.07 | (-0.88, 1.03) | 0.88 | 0.32 | (-0.40, 1.05) | 0.38 |
| Animal feces on floor where index child sleeps | Not visible | 58 | 1.52 | ref | | | | | |
| | Visible | 54 | 1.85 | 0.33 | (-0.05, 0.72) | 0.09 | 0.21 | (-0.12, 0.55) | 0.21 |
| Any animals on floor where index child sleeps | Not present | 71 | 1.67 | ref | | | | | |
| | Present | 41 | 1.70 | 0.02 | (-0.38, 0.43) | 0.91 | 0.19 | (-0.21, 0.58) | 0.36 |
| Chickens/ducks/pigeons nighttime stay | Outside home | 100 | 1.69 | ref | | | | | |
| | Inside home | 12 | 1.61 | -0.09 | (-0.81, 0.64) | 0.82 | 0.02 | (-0.64, 0.67) | 0.96 |
| Number of animals household owns | | | | | | | | | |
| Any animals | None | 25 | 1.55 | | | | | | |
| | 1st tertile | 33 | 1.9 | 0.35 | (-0.29, 0.99) | 0.28 | 0.06 | (-0.53, 0.66) | 0.84 |
| | 2nd tertile | 29 | 1.55 | 0 | (-0.62, 0.63) | 0.99 | -0.07 | (-0.64, 0.50) | 0.80 |
| | 3rd tertile | 25 | 1.68 | 0.14 | (-0.54, 0.81) | 0.69 | 0.10 | (-0.48, 0.67) | 0.74 |
| Cattle/buffalo | None | 61 | 1.5 | | | | | | |
| | 1st tertile | 25 | 1.71 | 0.21 | (-0.31, 0.73) | 0.44 | 0.13 | (-0.39, 0.64) | 0.63 |
| | 2nd tertile | 12 | 2.34 | 0.84 | (-0.1, 1.77) | 0.08 | **0.94** | **(0.05, 1.83)** | **0.04** |
| | 3rd tertile | 14 | 1.87 | 0.37 | (-0.37, 1.11) | 0.33 | 0.27 | (-0.31, 0.85) | 0.36 |
| Goats/sheep | None | 76 | 1.6 | | | | | | |
| | 1st tertile | 11 | 2.01 | 0.4 | (-0.46, 1.26) | 0.36 | -0.19 | (-1.03, 0.65) | 0.66 |
| | 2nd tertile | 14 | 1.95 | 0.35 | (-0.36, 1.06) | 0.33 | 0.05 | (-0.55, 0.64) | 0.88 |
| | 3rd tertile | 11 | 1.55 | -0.05 | (-0.85, 0.74) | 0.90 | -0.33 | (-0.88, 0.22) | 0.24 |
| Chickens/ducks/pigeons | None | 35 | 1.62 | | | | | | |

| | Exposure | N | Mean log10 CFU | Unadjusted | | | Adjusted | | |
|---|---|---|---|---|---|---|---|---|---|
| | | | | ΔLog10 | 95% CI | p-value | ΔLog10 | 95% CI | p-value |
| | 1st tertile | 26 | 1.83 | 0.21 | (-0.49, 0.91) | 0.55 | 0.05 | (-0.48, 0.57) | 0.86 |
| | 2nd tertile | 29 | 1.68 | 0.06 | (-0.51, 0.62) | 0.84 | 0.10 | (-0.42, 0.62) | 0.71 |
| | 3rd tertile | 22 | 1.6 | -0.02 | (-0.65, 0.61) | 0.95 | -0.01 | (-0.56, 0.53) | 0.96 |
| Number of animals compound owns | | | | | | | | | |
| Any animals | None | 16 | 1.74 | | | | | | |
| | 1st tertile | 35 | 1.62 | -0.12 | (-0.95, 0.7) | 0.77 | -0.53 | (-1.36, 0.29) | 0.21 |
| | 2nd tertile | 31 | 1.54 | -0.21 | (-0.93, 0.52) | 0.58 | -0.37 | (-1.03, 0.29) | 0.27 |
| | 3rd tertile | 30 | 1.88 | 0.14 | (-0.64, 0.92) | 0.73 | -0.15 | (-0.89, 0.58) | 0.68 |
| Cattle/buffalo | None | 51 | 1.47 | | | | | | |
| | 1st tertile | 23 | 1.54 | 0.06 | (-0.43, 0.55) | 0.81 | -0.17 | (-0.64, 0.30) | 0.47 |
| | 2nd tertile | 24 | 1.88 | 0.41 | (-0.18, 1) | 0.17 | 0.34 | (-0.24, 0.92) | 0.25 |
| | 3rd tertile | 14 | 2.34 | **0.86** | **(0.04, 1.69)** | **0.04** | **0.66** | **(0.08, 1.24)** | **0.02** |
| Goats/sheep | None | 65 | 1.58 | | | | | | |
| | 1st tertile | 21 | 1.81 | 0.23 | (-0.37, 0.83) | 0.46 | -0.26 | (-1.00, 0.48) | 0.49 |
| | 2nd tertile | 15 | 1.98 | 0.39 | (-0.37, 1.16) | 0.31 | 0.16 | (-0.49, 0.81) | 0.63 |
| | 3rd tertile | 11 | 1.61 | 0.02 | (-0.52, 0.57) | 0.94 | -0.23 | (-0.64, 0.18) | 0.26 |
| Chickens/ducks/pigeons | None | 23 | 1.83 | | | | | | |
| | 1st tertile | 32 | 1.57 | -0.26 | (-0.98, 0.47) | 0.49 | -0.53 | (-1.25, 0.18) | 0.14 |
| | 2nd tertile | 30 | 1.53 | -0.3 | (-0.89, 0.28) | 0.31 | -0.28 | (-0.91, 0.34) | 0.38 |
| | 3rd tertile | 27 | 1.86 | 0.04 | (-0.75, 0.82) | 0.93 | -0.12 | (-0.84, 0.61) | 0.75 |
| Animal roaming frequency inside home | | | | | | | | | |
| Any animals | 1st tertile | 36 | 1.41 | | | | | | |
| | 2nd tertile | 43 | 1.75 | 0.34 | (-0.16, 0.85) | 0.18 | 0.32 | (-0.17, 0.81) | 0.20 |
| | 3rd tertile | 33 | 1.89 | 0.49 | (-0.08, 1.06) | 0.09 | 0.20 | (-0.28, 0.68) | 0.40 |
| Cattle/buffalo | Never | 105 | 1.62 | ref | | | | | |
| | Sometimes | 7 | 2.61 | **0.98** | **(0.08, 1.89)** | **0.03** | **1.06** | **(0.27, 1.85)** | **0.009** |
| Goats/sheep | Never | 76 | 1.57 | ref | | | | | |
| | Sometimes | 31 | 1.97 | 0.40 | (-0.15, 0.96) | 0.15 | 0.12 | (-0.32, 0.56) | 0.59 |
| | Always | 5 | 1.56 | -0.01 | (-0.91, 0.89) | 0.99 | -0.17 | (-0.86, 0.53) | 0.64 |
| Chickens/ducks/pigeons | Never | 9 | 1.86 | ref | | | | | |
| | Sometimes | 32 | 1.44 | -0.41 | (-1.29, 0.46) | 0.35 | -0.42 | (-1.23, 0.38) | 0.30 |
| | Always | 71 | 1.77 | -0.09 | (-0.95, 0.77) | 0.84 | -0.26 | (-1.03, 0.50) | 0.50 |
| Animal roaming frequency in compound | | | | | | | | | |
| Any animals | 1st tertile | 58 | 1.71 | | | | | | |
| | 2nd tertile | 38 | 1.63 | -0.08 | (-0.58, 0.42) | 0.74 | -0.23 | (-0.73, 0.27) | 0.37 |
| | 3rd tertile | 16 | 1.7 | -0.01 | (-0.68, 0.66) | 0.97 | -0.27 | (-0.85, 0.31) | 0.36 |
| Cattle/buffalo | Never | 69 | 1.58 | ref | | | | | |
| | Sometimes | 33 | 1.88 | 0.30 | (-0.18, 0.78) | 0.22 | 0.02 | (-0.50, 0.53) | 0.94 |
| | Always | 10 | 1.74 | 0.16 | (-0.55, 0.87) | 0.66 | 0.22 | (-0.40, 0.85) | 0.49 |
| Goats/sheep roam | Never | 3 | 1.89 | ref | | | | | |
| | Sometimes | 6 | 2.29 | 0.41 | (-1.04, 1.85) | 0.58 | 0.93 | (-0.41, 2.26) | 0.17 |
| | Always | 103 | 1.64 | -0.25 | (-1.10, 0.61) | 0.58 | -0.20 | (-0.80, 0.39) | 0.50 |
| Chickens/ducks/pigeons | Never | 30 | 1.54 | ref | | | | | |
| | Sometimes | 53 | 1.92 | 0.38 | (-0.29, 1.04) | 0.27 | 0.02 | (-0.62, 0.67) | 0.94 |
| | Always | 29 | 1.39 | -0.15 | (-0.81, 0.50) | 0.64 | -0.39 | (-1.02, 0.25) | 0.23 |
| **Exposure domain 2: Antibiotic use in last 6 months** | | | | | | | | | |
| Index child (any antibiotic) | Did not use | 36 | 1.85 | ref | | | | | |
| | Used | 76 | 1.60 | -0.25 | (-0.71, 0.21) | 0.29 | -0.08 | (-0.54, 0.38) | 0.74 |
| Index child (beta-lactam) | Did not use | 83 | 1.62 | ref | | | | | |

| | Exposure | N | Mean log10 CFU | Unadjusted | | | Adjusted | | |
|---|---|---|---|---|---|---|---|---|---|
| | | | | ΔLog10 | 95% CI | p-value | ΔLog10 | 95% CI | p-value |
| | Used | 29 | 1.85 | 0.23 | (-0.29, 0.75) | 0.38 | 0.27 | (-0.17, 0.70) | 0.23 |
| Index mother (any antibiotic) | Did not use | 104 | 1.68 | | | | | | |
| | Used | 8 | 1.75 | 0.07 | (-1.02, 1.16) | 0.90 | -0.08 | (-1.10, 0.94) | 0.88 |
| Any animal in household | Did not use | 74 | 1.49 | ref | | | | | |
| | Used | 22 | 2.28 | **0.79** | **(0.23, 1.35)** | **0.006** | **0.63** | **(0.02, 1.24)** | **0.04** |

**Table S6.** Associations between log10-transformed colony-forming units (CFU) of ESBL-producing *E. coli* in **child stool, chicken feces, cow feces, courtyard soil, and floor swab** vs. number of animals in the household and compounds. The analyses controlled for potential socio-demographic confounders (child age and sex, mother's age and education, number of children <18 years in the household, number of individuals living in the compound, and asset-based wealth index) and study arm (intervention vs. control).

| Sample | Exposure | N | ΔLog10 (95% CI) [a] | p-value |
|---|---|---|---|---|
| Child stool | | | | |
| *Household level numbers of* | All animals | 113 | -0.01 (-0.18, 0.16) | 0.92 |
| | Chickens/ducks/pigeons | | 0.01 (-0.15, 0.17) | 0.90 |
| | Cattle/buffalo | | 0.07 (-0.04, 0.17) | 0.23 |
| | Goat/sheep | | **-0.24 (-0.45, -0.02)** | **0.03** |
| *Compound [b] level numbers of* | All animals | | 0.01 (-0.02, 0.05) | 0.37 |
| | Chickens/ducks/pigeons | | 0.01 (-0.02, 0.04) | 0.38 |
| | Cattle/buffalo | | **0.10 (0.01, 0.18)** | **0.02** |
| | Goat/sheep | | -0.06 (-0.20, 0.08) | 0.41 |
| Chicken feces | | | | |
| *Household level numbers of* | All animals | 65 | -0.19 (-0.54, 0.17) | 0.30 |
| | Chickens/ducks/pigeons | | -0.23 (-0.60, 0.14) | 0.22 |
| | Cattle/buffalo | | 0.03 (-0.06, 0.13) | 0.49 |
| | Goat/sheep | | -0.02 (-0.22, 0.19) | 0.87 |
| *Compound level numbers of* | All animals | | **0.06 (0.03, 0.09)** | **<0.005** |
| | Chickens/ducks/pigeons | | **0.06 (0.03, 0.09)** | **<0.005** |
| | Cattle/buffalo | | 0.08 (-0.01, 0.18) | 0.08 |
| | Goat/sheep | | 0.04 (-0.12, 0.19) | 0.66 |
| Cow feces | | | | |
| *Household level numbers of* | All animals | 62 | -0.16 (-0.55, 0.22) | 0.40 |
| | Chickens/ducks/pigeons | | -0.20 (-0.67, 0.28) | 0.41 |
| | Cattle/buffalo | | -0.05 (-0.14, 0.05) | 0.35 |
| | Goat/sheep | | -0.00 (-0.24, 0.24) | 0.97 |
| *Compound level numbers of* | All animals | | **0.07 (0.03, 0.11)** | **<0.005** |
| | Chickens/ducks/pigeons | | **0.07 (0.03, 0.11)** | **<0.005** |
| | Cattle/buffalo | | 0.02 (-0.09, 0.13) | 0.75 |
| | Goat/sheep | | 0.03 (-0.12, 0.17) | 0.7 |
| Courtyard soil | | | | |
| *Household level numbers of* | All animals | 112 | 0.15 (-0.05, 0.35) | 0.15 |
| | Chickens/ducks/pigeons | | 0.18 (-0.03, 0.38) | 0.09 |
| | Cattle/buffalo | | -0.02 (-0.12, 0.08) | 0.70 |
| | Goat/sheep | | 0.06 (-0.14, 0.26) | 0.55 |
| *Compound level numbers of* | All animals | | 0.01 (-0.02, 0.05) | 0.37 |
| | Chickens/ducks/pigeons | | 0.02 (-0.02, 0.05) | 0.36 |
| | Cattle/buffalo | | -0.01 (-0.12, 0.09) | 0.78 |
| | Goat/sheep | | 0.00 (-0.11, 0.11) | 0.97 |
| Floor swab | | | | |
| *Household level numbers of* | All animals | 112 | -0.00 (-0.18, 0.18) | 0.99 |
| | Chickens/ducks/pigeons | | -0.03 (-0.21, 0.14) | 0.73 |
| | Cattle/buffalo | | **0.09 (0.01, 0.17)** | **0.03** |
| | Goat/sheep | | -0.05 (-0.15, 0.04) | 0.28 |
| *Compound level numbers of* | All animals | | 0.01 (-0.01, 0.04) | 0.22 |
| | Chickens/ducks/pigeons | | 0.01 (-0.01, 0.04) | 0.27 |
| | Cattle/buffalo | | **0.08 (0.01, 0.15)** | **0.03** |
| | Goat/sheep | | -0.03 (-0.10, 0.04) | 0.44 |

CI: Confidence interval. Bolded estimates denote p-value<0.05.

[a] Δlog10-CFU associated with each additional 10 animals, 10 chickens/ducks/pigeons, 1 cattle/buffalo, 1 goat/sheep.

[b] Compound is a group of households around a central courtyard shared by extended families.
